# Humoral waning kinetics against SARS-CoV-2 is dictated by disease severity and vaccine platform

**DOI:** 10.1101/2024.10.17.24315607

**Authors:** Xin Tong, Benjamin Kellman, Maria-Jose Avendano, Maanasa Mendu, Jeff C. Hsiao, Eileen Serrano, Tamara Garcia-Salum, Nicolas Muena, Catalina Pardo-Roa, Mauricio Morales, Jorge Levican, Erick Salinas, Simone Cardenas-Cáceres, Arnoldo Riquelme, Nicole D. Tischler, Douglas A. Lauffenburger, Galit Alter, Ryan P. McNamara, Rafael A. Medina

## Abstract

SARS-CoV-2 vaccine-acquired immunity provides robust cross-variant recognition, while infection-acquired immunity can be heterogenous, with disease severity often modulating post-recovery responses. We assessed antibody waning dynamics between infection- and vaccination-acquired immunity across variants of concern (VOC). mRNA vaccination induced potent, cross-VOC Spike recognition and functional responses, but waned more rapidly for Omicron Spike. Hospitalized individuals developed more durable functional responses with lower peaks compared to mRNA vaccination, while outpatients exhibited slower decay than inactivated vaccine recipients. Humoral decay for the receptor binding domain tracked with neutralizing antibody titers, while S2-directed responses tracked with antibody-dependent myeloid cellular phagocytosis. Boosting the recovered patients with mRNA or inactivated vaccines expanded humoral breadth, durability, and restored functional responses, eliminating the severity- and platform-associated decay differences. Therefore, post-recovery hybrid immunization compensates for this distinction and broadens humoral breadth, highlighting the value of boosting immunity in previously infected individuals.

**One Sentence Summary:** Infection- and vaccine-acquired immunity to COVID-19 exhibit different functional antibody profiles, each characterized by distinct kinetics of waning over time.

## INTRODUCTION

As of August 2024, the severe acute respiratory syndrome coronavirus-2 (SARS-CoV-2) has infected over 676 million individuals, caused more than 6 million deaths, resulting in an unprecedented burden on healthcare systems and economies globally (*1–3*). Although vaccines against SARS-CoV-2 have proven highly successful in mitigating the global pandemic, emergent variants of concern (VOC) allowed the viruses to escape from the vaccine-associated antibody neutralization activities, which led to various levels of reduction in vaccine effectiveness since the beginning of the pandemic (*4–6*). In particular, the Omicron VOC lineage resulted in a significant increase in breakthrough infections globally (*7*). Although the SARS-CoV-2 vaccines are highly effective in preventing severe COVID-19 disease and death, the levels of neutralizing antibodies do not correlate entirely with protection against the SARS-CoV-2 vaccine breakthrough infection and reinfections, suggesting an important functional role of the vaccine-associated non-neutralizing immune functions in controlling the infection (*8–11*). More importantly, the durability of vaccine-induced immunity against VOCs remains poorly understood, which is vital in facilitating the development of updated SARS-CoV-2 vaccine candidates against future VOCs.

Previous studies have investigated the dynamics and durability of the vaccine-conferred protection and the levels of the neutralizing antibodies following natural SARS-CoV-2 infection or vaccination (*12, 13*). However, there is a limited consensus regarding the decay dynamics of neutralizing and non-neutralizing antibodies. While some studies demonstrated a rapid decline of neutralizing antibodies and protection months following viral exposure or vaccination (*12, 14, 15*), others reported a persisting level of neutralizing antibodies and, to a lesser extent, prolonged protection (*16, 17*). This discrepancy has been associated with other factors including the non-neutralizing functions of vaccine-induced antibodies. Beyond the conventional capability of antibodies to physically interact with pathogens, the binding antibodies can also recruit the innate immune components and trigger effector functions such as antibody-dependent phagocytosis (ADP) via interactions with the Fc-receptors expressed on all immune cells (*8, 18, 19*). Effector functions like ADP have been associated with a protective role against other viral infections (such as HIV-1 and CMV (*20*)). While the decay of neutralizing antibody titers between the different vaccine platforms has been exhaustively studied, the longitudinal decay dynamics of effector functions remain to be studied in the context of SARS-CoV-2 infection and vaccination response.

The Pfizer-BioNTech mRNA vaccine (BNT162b2) and Sinovac CoronaVac inactivated vaccines are two of the most widely distributed SARS-CoV-2 vaccines globally. Both vaccines exhibited strong protection against the original SARS-CoV-2 with BNT162b2 and CoronaVac exhibiting ∼95% and 84% effectiveness against severe illness, respectively (*3, 21, 22*). However, subsequent immunological studies reported a decline in neutralizing antibody titers in vaccinated individuals, which promoted the use of additional booster immunization (*23*). Although differences in antibody titers and neutralization afforded by these two vaccines have been documented (*24, 25*), it remains to be investigated whether the functional antibody responses induced by these two vaccine platforms decline differentially over time and whether heterologous boosting using these vaccines can promote or enhanced Fc functional immune signatures over pre-existing immunity acquired either by vaccination or prior infection.

Understanding immunity induced by exposure to the viral antigens through natural infection and immunization, known as “Hybrid Immunity” (*26*), has become of great interest since most of the population is now hybrid immune. Current data indicates that vaccination before or after SARS-CoV-2 infection can provide a more robust neutralizing immune response than either infection or vaccination alone (*27*). Here we focus on investigating the longitudinal dynamics and waning kinetics of functional humoral responses against wildtype SARS-CoV-2 and prototypic VOCs of infected (natural immunity) patients (outpatient and hospitalized) or individuals immunized with the BNT162b2 or CoronaVac vaccines (vaccine-induced immunity). We demonstrate that functional antibody responses against VOCs and their decay rates are associated with both disease severity and vaccine platform. In addition, given that most of the population have hybrid immunity against SARS-CoV-2 we analyze the effect of vaccine boosters after natural infection. We observed that the hybrid boost with the CoronaVac vaccine reversed the decay kinetics associated with having a severe infection and enhanced functional naïve-responses against Omicron. These results collectively demonstrate that the two vaccine platforms exhibited distinct functional antibody profiles and waning kinetics over time, and like heterologous boosting with an mRNA vaccine (*28*), the vaccination with inactivated virus after infection improves the humoral immune responses in terms of antibody levels and the breadth.

## RESULTS

### Vaccine platforms and COVID-19 clinical outcomes elicit subdomain-targeted antibodies with distinct decay kinetics

To explore longitudinal trends in immune response decay, we comprehensively profiled the humoral response in two cohorts: 1) vaccinated individuals (n = 49; overall median age: 33 years) with BNT162b2 (n = 15; median age: 36 years) or CoronaVac (n = 34; median age: 33 years) and 2) infected individuals (n = 77; overall median age: 38 years) managed as outpatients (n = 41; median age: 32 years) or hospitalized patients (n = 36; median age: 53.5 years) (**table S1**). A subgroup of the infected individuals were also subsequently vaccinated with either, the CoronaVac (Outpatient, n=26, median age: 33 years; or Hospitalized, n=16, median age: 53.5 year) or the BNT162b2 vaccines (Outpatient, n=3, median age: 27 years; or Hospitalized, n=2, median age: 55 years), and therefore these individuals were also assessed after acquiring hybrid immunity. Recruitment of infected patients was between March and December 2020, before the appearance of variants of concern (VOC).

While biphasic models are currently preferred for modeling antibody decay and estimating absolute parameters, these high-parameter models are inappropriate for high-throughput analysis. Hence, here we used low parameter single phase decay models and focused on relative comparisons between groups. The simplicity of these models enables high-throughput relative analysis without overfitting. Using this log-linear exponential decay approximation, which assumes a constant decay rate over time, we evaluated the half-life and initial values of circulating antibody titers using data from the peak of the immune response to ∼350 days post-infection or vaccination (see materials and methods). Initial IgG1 and IgG3 responses to WT, Delta, and Omicron BA.1 Spike were marked by differential magnitudes. mRNA vaccine recipient response was highest (BNT162b2 vaccine, dark violet line), followed by hospitalized recoverees (severe infection-acquired immunity, dark purple line), outpatient recoverees (mild infection-acquired immunity, light red line), and inactivated virus vaccine recipient responses (CoronaVac, light violet line) with the lowest initial responses (**Fig. 1A, top two rows**). IgA1 and IgM initial responses were highest in hospitalized individuals against WT and Delta Spike but were low-to-undetectable for all groups for Omicron BA.1 Spike (**Fig. 1A, bottom two rows**). mRNA-vaccine elicited IgG1 against the receptor binding domain (RBD) and full-length Spike and both had a half-life < 100 days, in contrast to IgG1 from hospitalized individuals, which showed a slower decay rate (**Fig. 1B, top row**). Similar IgG3 half-lives were observed for mRNA vaccination and hospitalization (**Fig. 1B second row**). While IgA1 and IgM showed a similar trend, only decay trends for hospitalized patients against wildtype and Delta were statistically significant (Wald-test FDR <0.05) for both the full Spike and RBD (**Fig. 1B, third and fourth rows**). In almost all cases, when decay models converged, outpatient recoverees and CoronaVac recipients showed lower initial response and nominally slower decay kinetics for full length Spike and RBD. These results demonstrate that initial antibody responses and sera longevity are associated differentially to distinct vaccine platforms and disease severity. Only hospitalized and outpatient recoverees exhibited nucleocapsid (N) reactive antibodies (**fig. S1a, data file S1**). While there were some associations between major sources of variance and select covariates, these had limited impact on the decay models. Scatter plots and antibody decay parameters are provided for all measured VOC Spike and RBD as well as all WT subdomains (**fig. S1-3**).

**Fig. 1.**
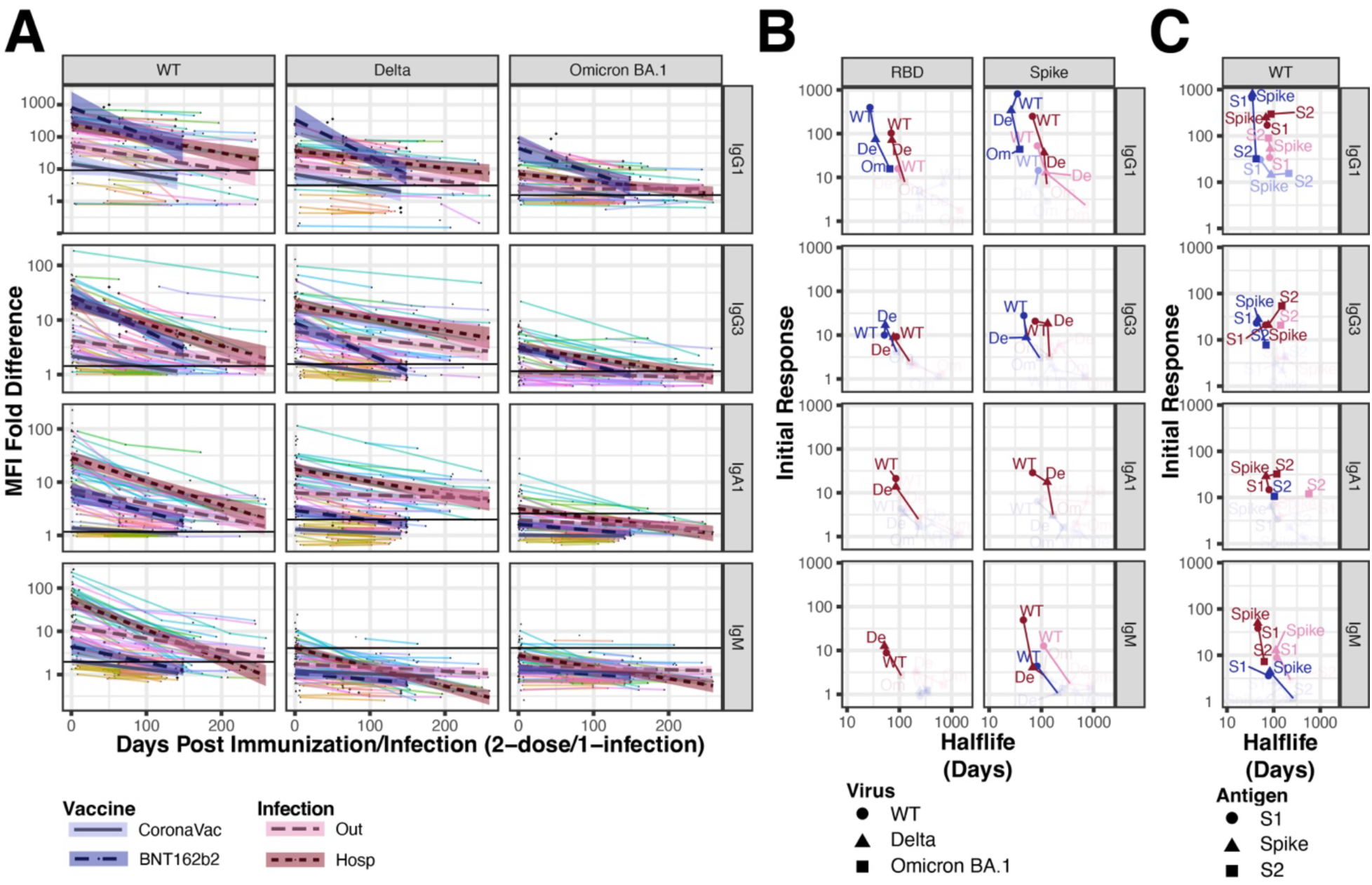
Antigen-specific decay over time post-infection or vaccination for binding immunoglobulins. (**A**) Scatter plots show subject-specific (line color) decline in immunoglobulin response specific to WT, Delta, or Omicron BA.1 Spike. Log-linear mixed-effect models with subject-specific random intercepts and slopes estimated trend-lines and 95% confidence intervals using data from Hospitalized (dark-purple) or Outpatient (light-pink) individuals, or subjects following the second BNT162b2 dose (dark-violet) or the second CoronaVac dose (light-violet). Horizontal black lines indicate the innate Spike reactivity in naïve (no-exposure) samples. Shown are the results for IgG1 (top row), IgG3 (second row), IgA1 (third row), and IgM (bottom row). The color legend is shown on the bottom. (B) Regression intercepts and slopes, indicating initial response and decay rate, are plotted with 95% confidence intervals for each variant of concern. Decay and response parameters across variants are also plotted together and connected within vaccination/infection group by lines. Legend for virus variant shape shown at the bottom; color scheme is like A. Shaded out regions indicate a response < the 97.5^th^ percentile of the naïve response. (C) Regression intercepts and slopes, indicating initial response and decay rate, are shown plotted with 95% confidence intervals for the two major Spike subdomains, S1 and S2. Legend for virus antigen shape shown on the bottom; the color scheme is like A. Shaded out regions indicate a response < the 97.5^th^ percentile of the naïve response. All units are median fluorescence intensity (MFI) normalized (fold difference) with respect to the naïve group.

Notably, both the response to an infection requiring hospitalization and the antibodies elicited by mRNA vaccination were substantial and remained detectable by our model between 200-400 days after the peak immune response (**fig. S4**), with IgG1, IgG3 and IgA1 antibody responses across VOC Spikes being the most durable for hospitalized patients. Additionally, although mRNA vaccine response decayed more quickly, the peak IgG1 response of mRNA-vaccinated individuals remained higher against Omicron BA.1 Spike than the response of hospitalized patients for nearly 171 days after the peak immune response (**Fig. 1A**).

Initial responses and decay kinetics were next examined for Fc-gamma receptor (FcγR) binding to antigen-reactive antibodies. Like the isotype-specific trends, FcγR showed high initial responses in mRNA vaccinees and COVID-19 hospitalized individuals, except for the responses against Omicron BA.1, where FcγR binding was diminished in hospitalized recoverees (**Fig. 2A).** Outpatient recoverees and CoronaVac recipients both showed lower initial FcγR-antibody interactions. Decay kinetics showed a similar trend to IgG1 and IgG3, with a longer half-life of FcγR-binding antibodies for hospitalized than for mRNA-vaccinated individuals **(Fig. 2B)**. While FcγR-binding antibodies against Omicron BA.1 Spike and RBD were lower, mRNA-vaccinated subjects antibody responses remained to have detectable responses for FcγRIIA and FcγRIIIA for nearly 150 days post-peak response. Additionally, FcγRIIA and FcγRIIIA binding of Omicron BA.1 specific antibodies remain higher in mRNA-vaccinated subjects than hospitalized patients for 105 and 116 days respectively (**Fig. 2A**).

**Fig. 2.**
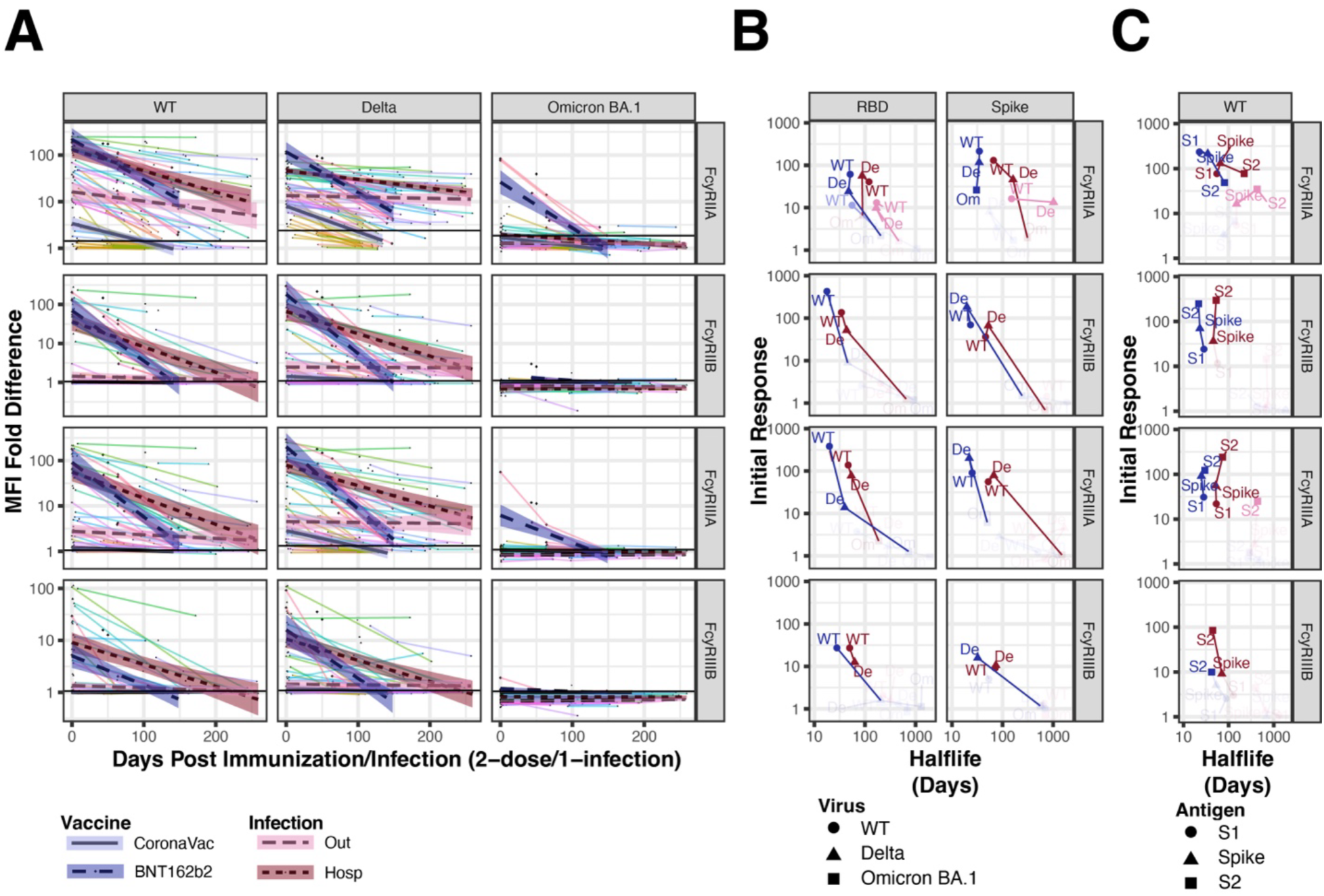
Antigen-specific decay over time post-infection or vaccination for FcR binding. **(A**) Scatter plots show subject-specific (line color) decline in FcγR binding specific to WT, Delta, or Omicron BA.1 Spike. Log-linear mixed-effect models with subject-specific random intercepts and slopes estimated trend-lines and 95% confidence intervals using data from Hospitalized (dark-purple), or Outpatient (light-pink) individuals, or subjects following the second BNT162b2 dose (dark-violet) or the second CoronaVac dose (light-violet). Horizontal black lines indicate the innate spike reactivity in Naïve (no-exposure) samples. Results are shown for FcγRIIA (top row), FcγRIIB (second row), FcγRIIIA (third row), and FcγRIIIB (bottom row). Color legend shown on bottom. (**B**) Regression intercepts and slopes, indicating initial response and decay rate are plotted with 95% confidence intervals for by each variant of concern. Decay and response parameters across variants are also plotted together and connected within vaccination/infection group by lines. Legend for virus variant shape shown on bottom; color scheme is like A. Shaded-out regions indicate a response < the 97.5^th^ percentile of the naïve response. (**C**) Regression intercepts and slopes, indicating initial response and decay rate, are shown plotted with 95% confidence intervals for the two major Spike subdomains, S1 and S2. Legend for virus antigen shape shown on the bottom; the color scheme is like A. Shaded-out regions indicate a response < the 97.5^th^ percentile of the naïve response. All units are median fluorescence intensity (MFI) normalized (fold difference) with respect to the naïve group.

To further subdivide the decay kinetics of SARS-CoV-2 Spike responses, we examined and compared the decay of antibodies specific for the S1 and S2 subunits of wild type Spike. IgG1 and FcγRIIA initial responses were primarily directed to the S1 subunit in mRNA vaccine recipients, whereas responses against S2 were predominant for the remaining FcγR-binding antibodies (**Fig. 1C and 2C**). In contrast, infected individuals produced a strong S2-directed initial response by IgG and IgA1 antibodies and most FcγR-binding antibodies (**Fig. 1C and 2C; fig. S1c**) with a longer decay rate. Notably, besides IgG1, CoronaVac recipients did not mount a significant response (>97^th^ percentile naïve response) against Spike subdomains using our thresholding approach. Taken together, mRNA- and infection-induced antibodies directed against S1 have high initial responses, but they also have the quickest decay rates. The more conserved S2 subdomain of Spike yielded more durable and functionally-primed antibody responses over time across groups, except for CoronaVac recipients.

### mRNA vaccinated subjects maintain a robust functional and durable response across VOCs

We next profiled functionality of the humoral response across our groups over time. Consistent with Fab and FcγR-binding antibodies, antibody-dependent cellular phagocytosis by monocytes (ADCP) and antibody-dependent neutrophil phagocytosis (ADNP) showed the highest initial response for mRNA-vaccinated and hospitalized individuals (**Fig. 3A, top row**). Functional decay of ADCP against WT Spike showed little decay over time for these two groups. CoronaVac recipients showed a similar initial ADCP response compared to hospitalized individuals but had a steeper rate of functional decline. Outpatient recoverees showed the lowest initial response, but ADCP was still detectable (>97^th^ percentile of naïve individuals) for close to 100 days. ADCP against Omicron BA.1 Spike showed a steeper rate of functional decay for BNT162b2 recipients, and a lower initial response for survivors of COVID-19 who required hospitalization. Interestingly, in the hospitalized group ADCP remained persistently detectable for 200 days. Neither CoronaVac recipients nor outpatient COVID-19 recoverees yielded an initial ADCP response significantly (Wald FDR > 0.05) above detectable levels for Omicron Spike (**Fig. 3A-B**, **top row**). Similar results were obtained for ADNP, with the exception that hospitalized individuals yielded no functional response above controls against Omicron Spike (**Fig. 3A-B**, **row 2**). Neutralization decays were also plotted for the four groups against WT SARS-CoV-2. Similar to binding assays to the RBD and S1, recipients of BNT162b2 and hospitalized recoverees mounted the highest initial neutralization to WT SARS-CoV-2 which waned to beneath the 97^th^ percentile cutoff within 50 and 100 days for BTN162b2 and hospitalized patients, respectively (**Fig. 3A, row 3**). Overall responses and decay kinetics to Omicron Spike were largely outside of the RBD, with the exception of IgG1 for BNT162b2 mRNA vaccine recipients. No other group showed a significant response and decay towards the Omicron RBD (**Fig.4**). Importantly, functional responses to Omicron BA.1 Spike remained present in the mRNA-vaccine recipients over time, providing support for the value of inducing sustained, non-neutralizing antibodies against SARS-CoV-2 Spike, capable of recognizing even highly divergent VOCs.

**Fig. 3.**
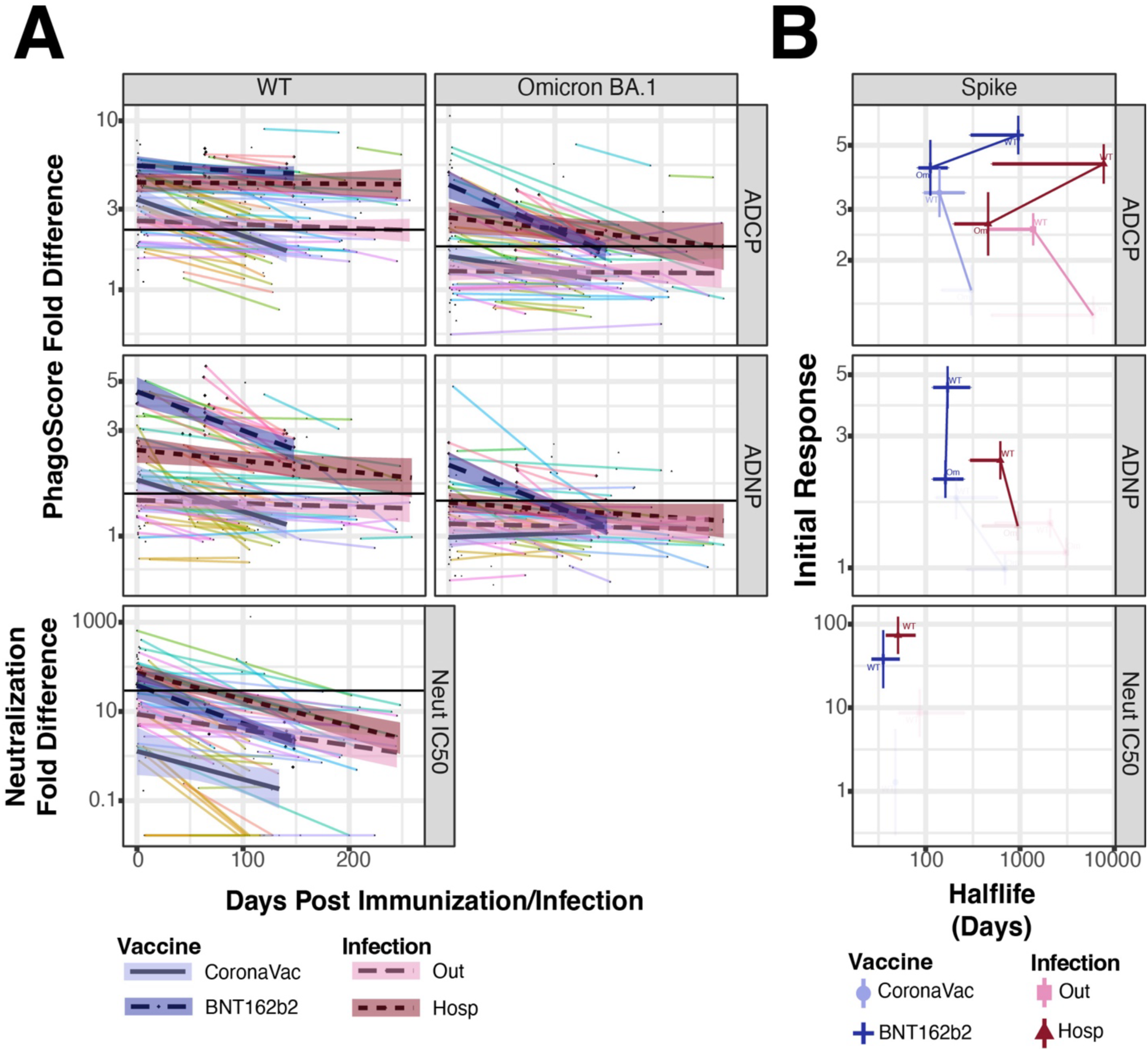
Functional Spike responses decay over time in a disease-severity and vaccine platform specific manner. (**A**) Scatter plots show subject-specific (line color) decline in phagocytotic functional response to WT and Omicron BA. 1 Spike. Log-linear mixed-effect models with subject-specific random intercepts and slopes estimated trend-lines and 95% confidence intervals using data from Hospitalized (dark-purple) or Outpatient (light-pink) individuals, or subjects following the second BNT162b2 dose (dark-violet) or the second CoronaVac dose (light-violet). Horizontal black lines indicate the innate spike reactivity in Naïve (no-exposure) samples. Shown are the antibody functional responses of ADCP (top row), ADNP (middle row), and neutralization (bottom row). Color legend shown on bottom. (**B**) Regression intercepts and slopes, indicating initial response and decay rate, are plotted with 95% confidence intervals for WT and Omicron BA.1 Spike. Decay and response parameters across variants are stratified by infection/vaccination type. Legend for virus variant shape shown on bottom; color scheme is like A. Shaded out regions indicate a response < the 97.5^th^ percentile of the naïve response.

**Fig. 4.**
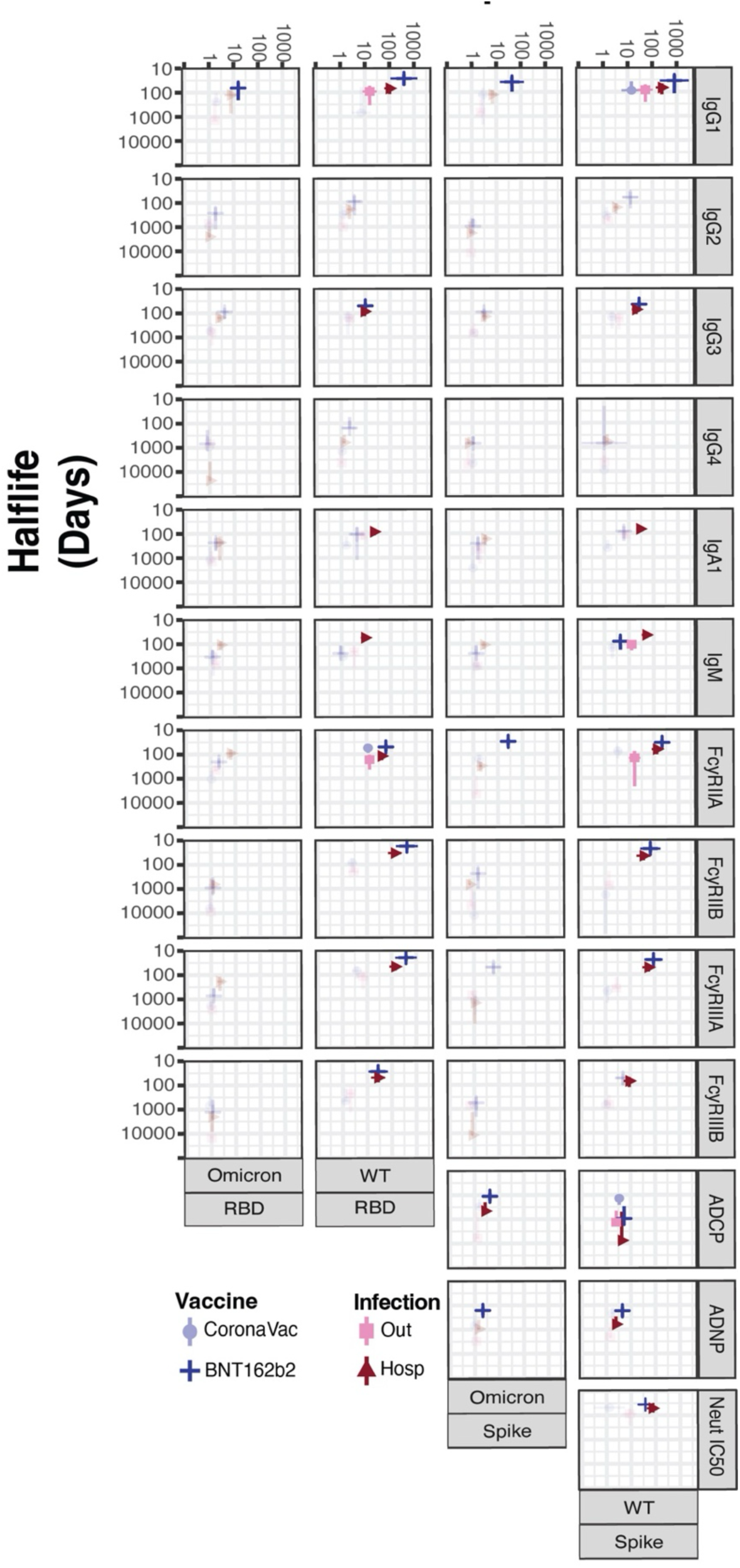
Overall responses to Omicron Spike are largely outside of RBD. Decay parameters are shown for WT and Omicron BA.1 Spike across all assays. Color and shape scheme is like A and B. Shaded out regions indicate a response < the 97.5^th^ percentile of the naïve response and a naïve-fold-difference of 2. X-axis units are MFI fold difference for all antibody binding assays, Phagoscore fold difference for ADCP and ADNP, and IC50 fold difference for neutralization.

### Hybrid boosting rescues decayed responses and eliminates severity-dependent responses

We next examined if infected individuals who received either the inactivated CoronaVac or BNT162b2 mRNA vaccines after recovery (hybrid immunity) mounted/restored a functional humoral response to WT and Omicron Spike. CoronaVac recipients who previously had outpatient COVID-19 illness had a significant boost in IgG1, FcγRIIA and FcγRIIIA towards both WT and Omicron Spike (**Fig. 5A-B**). Hospitalized COVID-19 patients only had a significant (paired t-test FDR < 0.05) boost to Omicron Spike for FcγRIIIA-binding antibodies, while other antibodies trended positivity, but mostly failed to reach statistical significance (**Fig. 5A-B; fig.S5a-b**). Individuals who received the BNT162b2 vaccine post-recovery also showed enhanced humoral responses, but only FcγRIIA and FcγRIIIA to WT Spike and FcγRIIIA to Omicron BA.1 Spike yielded statistically significant increases after multiple test corrections. This could be due to the limited number of individuals who received mRNA vaccines.

**Fig. 5.**
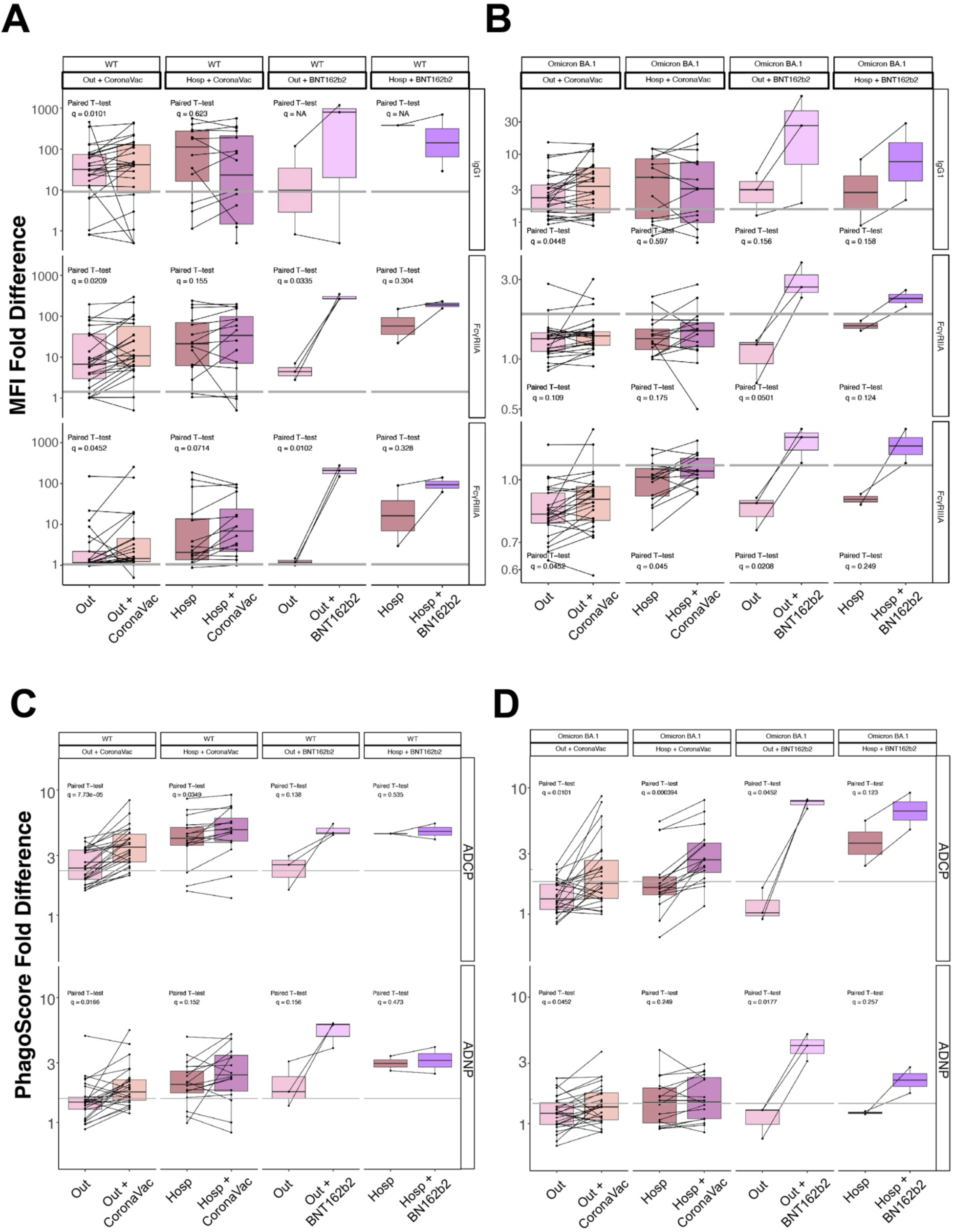
Hybrid immune response of vaccinated individuals post SARS-CoV-2 infection. Paired t-tests are visualized for Omicron and WT Spike binding (**A, B**) and function (**C, D**) for naturally infected individuals (hospitalized or outpatients) immunized with either the CoronaVac or BNT162b2 vaccines post-infection. The infection-only boxplots (Out or Hosp) represent the fully waned response (response at the farthest timepoint following infection) while the paired post-infection vaccination boxplots represent peak immunity following vaccination. The grey line indicates the 97th percentile of naïve response. Q-values indicate the FDR corrected significance for each corresponding paired t-test. Y-axis units for A and B are MFI fold difference, while Y-axis units for C and D are Phagoscore fold difference.

ADCP in previously infected patients who subsequently received CoronaVac also showed highly significant increases regardless of disease severity for both WT and Omicron Spike. In contrast, only outpatient recoverees showed a significant increase in ADNP for WT and Omicron Spike after CoronaVac vaccination (**Fig. 5C-D**). For recipients of BNT162b2 post-recovery, a significant increase in ADCP and ADNP was only observed for Omicron Spike in the outpatient group. Again, this could be attributable to a low sample in this hybrid group. Binding antibodies and FcγR-binding antibodies to Spike subdomains (**fig. S6a**) and VOCs (**fig. S6b**) for these hybrid groups also showed considerable humoral breadth.

We noted that hybrid boosting with the inactivated CoronaVac vaccine appeared to remove the severity-dependent response distinctions in some humoral responses. Thus, we directly compared how hybrid boosting with CoronaVac affected humoral profiles relative to how they were initially shaped by disease severity. Hospitalized recoverees (dark purple) had significantly higher initial IgG3, FcγRIIA- and FcγRIIIA-binding antibodies to WT Spike compared to outpatient recoverees; however, this significant separation was eliminated through subsequent vaccination with CoronaVac (hybrid immunity; **Fig. 6A, first to third rows)**. Similarly, ADNP was significantly higher for hospitalized recoverees, but hybrid immunity conferred by CoronaVac vaccination eliminated this discrepancy (**Fig. 6A, bottom row**). Similar trends were also observed for Omicron BA.1 Spike, albeit with lower initial and subsequent hybrid responses. The notable exception was FcγRIIIA binding to Omicron Spike (**Fig. 6B**). Other initial differences between outpatient and hospitalized COVID-19 recoverees were also narrowed or completed ablated post-vaccination, indicating that the functional breadth of hybrid immunity was largely disease-severity independent (**fig. S7 and S8**).

**Fig. 6.**
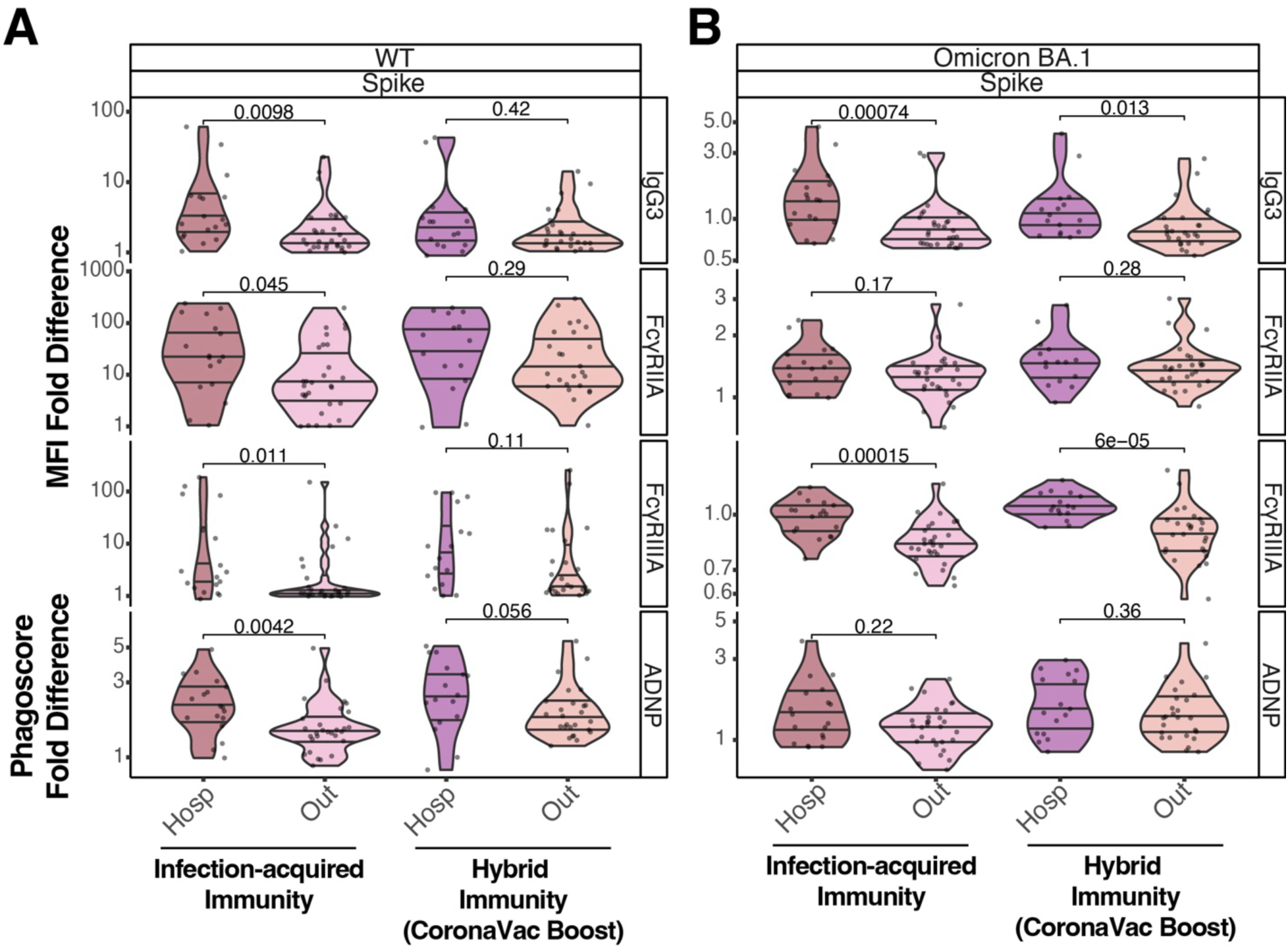
Hybrid immunity induced by inactivated, or mRNA vaccination rescues the disease-severity dependent humoral decay. (A) Violin plots showing waned post-infection (left) to peak response following the post-infection vaccination with CoronaVac (right). Shown are responses for WT Spike for IgG3 (top row), FcγRIIA- (second row) and FcγRIIIA-binding antibodies (third row), and ADNP (bottom row). Q-values are shown above pairwise comparisons for each subgroup. (B) Same as A, but for responses to Omicron BA.1 Spike. Y-axis units are MFI fold difference and Phagoscore fold difference.

## DISCUSSION

Antibodies play a critical role in the control of SARS-CoV-2 infection and the development of severe COVID-19. Numerous studies have shown that vaccine platform and disease severity result in distinct humoral responses against SARS-CoV-2. While neutralizing antibody levels are known to decay over time (*29–34*), limited work has been done to characterize non-neutralizing antibody waning (*28, 35*). Characterizing these decay kinetics is critical to understanding functional immune responses and the influence of disease severity and vaccine platforms on antibody responses. Here we show that the initial response and half-life of both antibody and Fc effector functions depend on the vaccine platform and disease severity. The observed differences in immune response between mRNA vaccination and SARS-CoV-2 infection reported here are consistent with other reports, where vaccination elicited a more robust response that decays more rapidly over time compared with natural infection (*36*), which is further influenced by VOC. Notably, hybrid boosting -vaccination of a COVID-19 recoveree-narrowed or eliminated the severity-dependent decay dynamics. We also found that most antibodies directed against the S2 domain of SARS-CoV-2 Spike displayed the longest half-life compared to antibodies directed against other regions of Spike, which could be attributed to previous exposure to endemic human coronaviruses (*37, 38*). In contrast, antibodies directed towards the RBD domain, which is the most immunogenic region and comprise of the majority of Spike-specific neutralizing activity, have a shorter half-life. This is consistent with prior works that have established that RBD alone is sufficient to capture the majority of neutralization effects and decay dynamics against SARS-CoV-2 (*31*).

We observed that the effector functions of Fc generated by natural infection were not affected in the same way as the vaccine-generated immunity against VOCs. ADCP maintained cross-reactivity between recovered individuals requiring hospitalization and those immunized with the mRNA vaccine; by contrast, only ADNP cross-reactivity was observed against Omicron in mRNA vaccinees. Interestingly, both functional responses induced by the mRNA vaccine were directed to less divergent epitopes of the protein, which agree with those described for other non-neutralizing effector functions such as ADCC (*39–41*). Furthermore, this targeted response to more conserved regions of the Spike protein, such as the S2 region, could be associated with the slower decay kinetics of the ADCP response against Spike WT observed in both groups. However, this would not account for the kinetics of ADNP decay in vaccinees and the cross-reactivity of this function in infected individuals, which in concordance with other studies, suggest that not all Fc effector functions have their target in the S2 subunit or outside the RBD domain (*39, 42*).

We also examined the disease severity and vaccine platform impact on immune response durability over viral evolution—across VOCs. For most measures, particularly FcR binding and functional response, initial response and half-life decreased dramatically in response to Omicron BA.1 antigens. Only mRNA-vaccinated individuals showed a non-naïve FcyRIIA, and FcyRIIIA initial response to Omicron spike. Even in these subjects, FcR binding and functional response remained above non-naïve levels only briefly.

While this study is not intended to evaluate the performance of the different immunization platforms, we note that, like hybrid immunization with mRNA platforms, post COVID-19 boosting with the inactivated virus platform provided improvements in multiple measures of the immune response to Omicron Spike (*27, 37, 43*). IgG1 binding, FcyRIIIA, ADCP, and ADNP, increased significantly following hybrid immunization with CoronaVac. In various measures that had waned below naïve response, these responses were recovered. This supports and highlights the importance of vaccination of previously infected individuals given that most antibody activity wanes below detectable levels in approximately 200 days after COVID-19. Importantly, beyond the recovery of these measures, we observed the elimination of the severity-dependent response between outpatient and hospitalized patients following immunization. This distinct feature seen in infected-immunized individuals (hybrid immunity) resembles the extinction of the platform-specific effects described following heterologous boosting (*44*). Although we recognize that biphasic models are currently preferred for modeling antibody decay, we decided to use the low parameter single phase decay models due to its suitability for our high-throughput relative analyses. However, a limitation of this approach is that we did not evaluate the fit of monophase or biphasic decay to these parameters, possibly resulting in less accurate half-life calculations. Another limitation of this study refers to the opportunistic sampling of the study cohort. As a result, our study design was not powered to account for some possible correlations with covariates (e.g. age, clinical features and comorbidities). Such associations might have an impact on the decay dynamics and therefore, further studies should be performed to further established the role of covariates in antibody activity, function and longevity.

Collectively our results demonstrate that waning kinetics of non-neutralizing humoral responses are associated with vaccine platform and disease severity, across subdomains of the Spike protein. Thes data help to establish a rational for developing future vaccines and boosting strategies against SARS-CoV-2.

## MATERIALS AND METHODS

### Study design and Subject Details

An informed written consent form was obtained under protocol 200829003, which was reviewed and approved by the Ethics Committee at Pontificia Universidad Católica de Chile (PUC). This work was also supervised and approved by the Emory University Institutional Review Board (IRB ID STUDY00005857) and the Mass General Institutional Review Board (IRB #2020P00955 and #2021P002628).

Serum samples were obtained from two cohorts: 1) Subjects who received the complete-dosage regimen for the respective vaccines recommended by the manufacturers. The vaccine cohort contained samples from individuals who received either the mRNA BNT162b2 (n = 15; median age: 36 years [range 15–53] 3% male) or the inactivated CoronaVac vaccines (n = 34; median age: 33 years [range 21–80] 10% male). The BNT162b2 vaccine group was given 30 μg BNT162b2 (mRNA vaccine encoding the Spike of SARS-CoV-2 Wuhan-HU-1 strain) on days 0 and 21 (**Table S1**). The CoronaVac group received two doses of 600 U CoronaVac (inactivated SARS-CoV-2 Virus (CZ02 strain) four weeks apart. 2) Infected individuals who required hospitalization (n = 36, median age: 53.5 years [range 16–83], 25 % male) or were outpatients (n = 41, median age: 32 years [range 13–66], 21% male). A subgroup of infected individuals who received either CoronaVac (Outpatient, n=26, median age: 33 years [range 18-66] 11% male; or hospitalized, n=16, median age: 53.5 years [range 29-83] 10% male) or the BNT162b2 vaccines (Outpatient, n=3, median age: 27 years [range 13-30] 1% male; Hospitalized, n=2, median age: 55 years [range 48-62] 1% male).

### Antigens

All antigens used in this study are listed in **Table S2**. Most antigens were lyophilized powder and were resuspended in water to afford a final concentration of 0.5 mg/mL. Antigens that required biotinylation were treated with the NHS-Sulfo-LC-LC kit per the manufacturer’s instruction. Excess biotin and buffer exchange from Tris-containing antigens were removed using the Zebra-Spin desalting and size exclusion chromatography columns.

### Immunoglobulin Isotype and Fc Receptor Binding

Antigen-specific antibody levels of isotypes and subclasses and levels of Fcγ-receptor binding were evaluated using a custom multiplexing Luminex-based assay platform in technical replicates, as previously described (34). The antigens were directly coupled to magnetic Luminex beads (Luminex Corp, TX, USA) by carbodiimide-NHS ester-coupling chemistry, which designates each region to each antigen. Individual dilution curves for each antigen were performed to identify an appropriate dilution factor for each secondary feature within the linear range of detection. The antigen-coupled beads were incubated with different serum dilutions (1:100 for IgG2, IgG3, IgG4, IgM, and IgA1, 1:500 for IgG1, and 1:750 for Fcγ-receptor binding) overnight at 4°C in 384 well plates (Greiner Bio-One, Germany). Unbound antibodies were removed by washing and subclasses, isotypes were detected using the respective PE-conjugated antibody listed in Supplementary Table 2. All detection antibodies were used at a 1:100 dilution.

For the analysis of Fcγ-receptor binding PE-Streptavidin (Agilent Technologies, CA, USA) was coupled to recombinant and biotinylated human FcγRIIA, FcγRIIB, FcγRIIIAV, or FcγRIIIB protein. Coupled Fcγ-receptors were used as a secondary probe at a 1:1000 dilution. After 1 hour of incubation, the excessive secondary reagent was removed by washing, and the relative antibody concentration per antigen was determined on an IQue Screener PLUS cytometer (IntelliCyt).

### Evaluation of Antibody-mediated Functions

A flow cytometry-based phagocytic assay was used to evaluate antibody-dependent cellular phagocytosis (ADCP) and neutrophil phagocytosis (ADNP) using fluorescently labeled microspheres, as described previously (*45*). The listed WT and Omicron Spike antigens were biotinylated and conjugated to yellow-green, fluorescent neutravidin microspheres. Then diluted serum samples with the pre-determined concentrations (1:100) were incubated with the coupled antigens. The pre-formed immune complexes bound with microspheres were washed and incubated with a human monocyte cell line (THP-1) for ADCP function or with neutrophils collected from healthy donors’ blood samples to assess ADNP activity. Cells studied under ADNP assays were then stained with anti-CD66b Pac blue antibody to calculate the percentage of CD66b+ neutrophils. In both ADCP and ADNP assays cells were fixed with 4% paraformaldehyde (PFA) and identified by gating on single cells and microsphere-positive cells. Microsphere uptake was quantified as a phagocytosis score, calculated as the (percentage of microsphere-positive cells) x (MFI of microsphere-positive cells) divided by 100000.

### Microneutralization Assay using rVSV SARS-CoV-2 Spike Protein (rVSV-SARS2-S)

To determine the neutralization antibody titers of patient sera, we used a previously described the replication-competent recombinant vesicular stomatitis virus carrying the SARS-COV2 spike protein and coding for an enhanced green fluorescent protein (eGFP) (*46*). As described previously (*29*), Vero E6 cells grown in 1X MEM supplemented with 10% FBS were transfected with plasmid pCEP4-myc-ACE2 and the stable clones were selected by hygromycin (400 mg/mL). To assay neutralization antibody titers, serial dilutions of serum samples were incubated with rVSVSARS2-S for 1 h at 37 °C. The serum-virus inoculum was added to Vero E6 hACE2 cells seeded the day before in optical bottom 96-well plates (Thermo Scientific) at 80% confluence and adsorbed for 2 h at 37 ° C. Next, the mixture was replaced by culture media and infection allowed to proceed for 20 h at 37 °C, 5% CO2 and 80% humidity. The cells were then fixed with 4% formaldehyde (Pierce) and stained in with 40, 6-diamidino-2-phenylindole (DAPI) 300 nM (Invitrogen). Viral infectivity was quantified by automated enumeration of GFP-positive cells (normalizing against cells stained with DAPI) using a Cytation5 automated fluorescence microscope (BioTek) and segmentation algorithms applied from the ImageJ program. Alternatively, total GFP fluorescence per well was acquired using the Cytation5 fluorescence lector (wavelength for DAPI 360 nm for absorption, 460 nm for emission and for GFP, 485 nm for absorption, 526 nm for emission) and normalized against DAPI fluorescence. The half maximum inhibitory concentration (IC50) of the sera, were calculated from data obtained with two technical replicates using non-linear regression analysis and the curve fitting was done using second-order polynomial (quadratic); and linear regression models (using log10 IC50 transformed data) were done with GraphPad Prism 5 software.

### Quantification and Statistical Analysis

All data analysis was done using R Studio 1.4.1103 or FlowJo. Statistical analysis was done using R studio or GraphPad Prism. No data point was omitted from the analysis. Box and whisker plots were generated using ggplot, calculating the mean and standard deviation for each factor. Technical replicates for each sample were performed and the mean between replicates was plotted. Coefficient of Variance (CV) was measured for each measurement (replicate variance / replicate mean for each sample, antigen and detector combination). High CV measurements (CV>30% for isotype/FcR measurements, CV>50% for functional measurements) were dropped.

### Antibody Decay Estimation

Data were background corrected by subtracting the median MFI for PBS controls on each plate. All MFI less than 1000 were excluded as null values. We compared subjects following their second vaccination or their first infection. Samples before the peak immune response (those followed by a higher measurement) were excluded.

The decay rates and initial values of antigen-specific antibody titers was estimated using a log-linear approximation of exponential decay. Using Rv3.1 (Lindstrom and Bates 1990), we fit univariate temporal mixed effect model predicting log (MFI) from days post-infection/inoculation with random intercepts and slopes for each subject. To mitigate processive noise we used a first-order autoregressive correlation structure. One model was fit for antibody titers specific to each VOC spike antigen and each immunization/infection-type. Initial response and decay Half-life correspond to the exponentiated intercept and log (2)/slope of the log-linear decay models. Confidence intervals (95%) were calculated for initial response and half-life using standard errors fit in the regressions. Regression confidence intervals (95%) were calculated using a point-wise confidence interval (Wickham 2016).

To determine the time (t) to convergence between two groups (α and β), we set the regression formulas equal at:

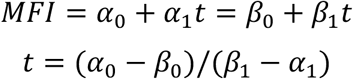

We asserted a zero-slope for comparison to naïve samples. Confidence intervals (95%) were calculated by scaling the projection by standard error (SE) for each regression parameter estimate (E)

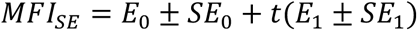

## Data Availability

All data produced in the present work are contained in the manuscript.

https://github.com/bkellman/COVID_functional_decay_2024

## Supplementary Materials

Figs. S1 to S7

Supplementary Fig 1: WT spike subdomain-specific MFI decay over time post-infection or vaccination for Immunoglobulin and FcR

Supplementary Fig 2: Spike-specific MFI decay across VOC over time post-infection or vaccination for Immunoglobulin and FcR

Supplementary Fig 3: RBD-specific MFI decay across VOC over time post-infection or vaccination for Immunoglobulin and FcR

Supplementary Fig 4: Time to decay calculated from the log-linear regression models.

Supplementary Fig 5: Comparison of the peak response post-infection to the peak response following post-infection across VOC for Immunoglobulin and FcR.

Supplementary Fig 6: Hybrid boosting expands binding breadth for both outpatient and hospitalized recoverees.

Supplementary Fig 7: Comparison of the peak response post-infection to the peak response following post-infection.

Supplementary Fig 8: Comparison of waning with peak response following post-infection boosting.

Tables S1 to S2

Supplementary Table 1: Demographic and baseline characteristics.

Supplementary Table 2: List of reagents and resources used in this study.

Data file S1: https://github.com/bkellman/COVID_functional_decay_2024/blob/main/data-final/DataSet1%20-%20DS04-data_decay.rand_intercepts.just_model.csv

## Acknowledgments

The authors would like to thank the laboratory of Prof. Douglas Lauffenburger (Massachusetts Institute of Technology) for the critical evaluation of statistics in this manuscript. We also thank Mark and Lisa Schwartz, Terry and Susan Ragon, and the SAMANA Kay MGH Research Scholars award for their support. We also would like to thank Estefany Poblete for her technical assistance during sample processing.

## Funding

Massachusetts Consortium on Pathogen Readiness (MassCPR) (G.A. and R.P.M.)

Gates Global Health Vaccine Accelerator Platform INV-001650 (G.A. and R.P.M.)

NIH (2U19AI135995-06, U19AI42790-05, 1P01AI168347-01, P01AI165072-1, U01CA260476-0251) (G.A. and R.P.M.)

FLUOMICS and SYBIL Consortium (NIH-NIAID grant U19AI135972) (RAM)

Center for Research on Influenza Pathogenesis (CRIP), an NIAID Center of Excellence for

Influenza Research and Surveillance (CEIRS, contract # HHSN272201400008C) (R.A.M.)

FONDECYT 1212023 grant from ANID of Chile R.A.M.)

FONDECYT 1221811 grant from ANID of Chile (N.D.T.)

FONDECYT 11231122 grant from ANID of Chile (C.P.R.)

Centro Ciencia & Vida, CCTE Basal FB210008 (N.D.T.)

Postdoctoral grant FONDECYT 3190706 (C.P.R.)

Postdoctoral grant FONDECYT 3190648 (J.L.)

ANID Becas/Doctorado Nacional 21212258 (M.J.A.)

Scholarship from Vicerrectoria de Investigacion de la Escuela de Graduados, Pontificia Universidad Catolica de Chile. (E.S.)

## Author Contributions

Conceptualization: X.T., BK., M.J.A., R.P.M., and R.A.M.

Methodology: X.T., BK., R.P.M., G.A, M.J.A., N.M., S.C., N.T. and R.A.M.

Investigation: X.T., BK., M.J.A., R.P.M., and R.A.M.

Validation: X.T., BK., M.J.A., R.P.M., and R.A.M.

Formal Analysis: X.T., BK., M.J.A., M.M., R.P.M., and R.A.M.

Cohort study design, supervised and managed the sample collection: E.F.S., T.G.S., C.P.R., A.R., and R.A.M.

Processed samples, revised the manuscript: E.F.S., T.G.S., C.P.R., J.L., E.S., A.R., M.J.A., N.M., S.C., N.T. and R.A.M.

Resources: G.A., and R.A.M.

Writing Original Draft: X.T., BK., and M.J.A.

Writing Review & Editing: R.P.M., G.A., and R.A.M.

Visualization: B.K.

Project Administration: R.P.M., and R.A.M.

Funding Acquisition: G.A., N.T. and R.A.M.

Supervision, R.P.M., R.A.M. and G.A.

## Corresponding Authors

Correspondence to Ryan P. McNamara or R. A. Medina.

## Data and Materials Availability

### Data Availability

All the data generated in this study are provided within the article and the Supplementary data is provided as a Source Data file. The Systems Serology data generated in this study are available in the GitHub database: https://github.com/bkellman/COVID_functional_decay_2024.

### Code Availability

All coding was done using R Studio V 1.4.1103 using ggplot. Individual groups were analyzed as factors. All codes and scripts are available upon request to the lead data point of contact. This article does not report original code. Any additional information required to reanalyze the data reported in this article is available from the lead contact upon request.

