## Supplementary Tables and Figures for "Humoral waning kinetics against SARS-CoV-2 is dictated by disease severity and vaccine platform"

**Supplementary Material**

Contains Tables S1-S2

Contains Figs S1-S8 and corresponding figure captions

**Supplementary Table S1. Demographic and baseline characteristics of the cohort.**

| | Outpatients (a) (n = 41) | Hospitalized (b) (n = 36) | CoronaVac (c) (n = 34) | BNT162b2 (d) (n = 15) | P value* | P value# | Outpatients CoronaVac (n = 26) | Hospitalized CoronaVac (n = 16) | P value^ | Outpatients BNT162b2 (n = 3) | Hospitalized BNT162b2 (n = 2) | P value^ |
|---|---|---|---|---|---|---|---|---|---|---|---|---|
| ***Characteristics*** | | | | | | | | | | | | |
| Male, n (%) | 21 (51.2) | 25 (69.4) | 10 (29.4) | 3 (20.0) | **0.0010** | **0.010 (b,c)**<br>**0.011 (b,d)** | 11 (42.3) | 10 (62.5) | 0.3408 | 1 (33.3) | 1 (50) | 1 |
| Age, median (IQR) | 32<br>(40-26) | 53.5<br>(62.5-37.0) | 33<br>(44-27.25) | 36<br>(38.5-24.5) | **< 0.0001** | **0.0002 (a,b)**<br>**0.0009 (b,c)**<br>**0.0040 (b,d)** | 33<br>(54.75-27.75) | 53.5<br>(62.5-37) | **0.0295** | 27<br>(28.5-20) | 55<br>(58.5-51-5) | 0.1489 |
| > 60 years, n (%) | 4 (9.8) | 12 (33.3) | 1 (2.9) | 0 (0) | **0.0007** | **0.0083 (b,c)** | 3 (11.5) | 5 (31.3) | 0.2233 | 0 (0) | 1 (50) | 0.4 |
| ***Comorbidities or conditions*** | | | | | | | | | | | | |
| Obesity (BMI ≥ 30), n (%) | 5 (12.2) | 13 (36.1) | 5 (14.7) | 3 (20) | 0.0613 | NS | 2 (7.7) | 7 (43.8) | **0.0161** | 1 (33.3) | 1 (50) | 1 |
| Hypertension, n (%) | 3 (7.3) | 12 (33.3) | 3 (8.8) | 2 (13.3) | **0.0119** | **0.047 (a,b)** | 3 (11.5) | 6 (37,5) | 0.0628 | 0 (0) | 0 (0) | NA |
| Cardiovascular disease, n (%) | 0 (0) | 3 (8.3) | 0 (0) | 0 (0) | 0.0774 | NS | 0 (0) | 1 (6.3) | 0.3810 | 0 (0) | 0 (0) | NA |
| Chronic pulmonary disease, n (%) | 6 (14.6) | 3 (8.3) | 0 (0) | 0 (0) | 0.0623 | NS | 4 (15.4) | 1 (6.3) | 0.6332 | 1 (33.3) | 0 (0) | 1 |
| Asthma, n (%) | 8 (19.5) | 2 (5.6) | 4 (11.8) | 4 (26.7) | 0.1310 | NS | 6 (23.1) | 0 (0) | 0.0671 | 1 (33.3) | 0 (0) | 1 |
| Rheumatologic disease, n (%) | 0 (0) | 2 (5.6) | 0 (0) | 1 (6.7) | 0.1327 | NS | 0 (0) | 1 (6.3) | 0.3810 | 0 (0) | 0 (0) | NA |
| Immunocompromise, n (%) | 0 (0) | 5 (13.9) | 0 (0) | 0 (0) | **0.0069** | NS | 0 (0) | 3 (18.8) | 0.0488 | 0 (0) | 0 (0) | NA |
| Allergy+, n (%) | 20 (48.8) | 6 (16.7) | 17 (50) | 6 (40) | **0.0089** | **0.023 (a,b)**<br>**0.028 (b,c)** | 12 (46.2) | 2 (12.5) | **0.0420** | 3 (100) | 0 (0) | 0.1 |
| Neurologic disease, n (%) | 0 (0) | 4 (11.1) | 0 (0) | 0 (0) | **0.0217** | NS | 0 (0) | 2 (12.5) | 0.1394 | 0 (0) | 0 (0) | NA |
| Smoker, n (%) | 6 (14.6) | 9 (25.0) | 5 (14.7) | 4 (26.7) | 0.4962 | NS | 4 (15.4) | 4 (25) | 0.4538 | 1 (33.3) | 0 (0) | 1 |
| ***Previous treatment*** | | | | | | | | | | | | |
| Immunosuppressive drugs, n (%) | 0 (0) | 5 (13.9) | 0 (0) | 0 (0) | **0.0071** | NS | 0 (0) | 3 (18.8) | **0.0488** | 0 (0) | 0 (0) | NA |
| ***Symptoms*** | | | | | | | | | | | | |
| ***Respiratory*** | | | | | | | | | | | | |
| Cough, n (%) | 28 (68.3) | 30 (83.3) | NA | NA | 0.1855 | | 19 (73.1) | 13 (81.3) | 0.7152 | 3 (100) | 2 (100) | NA |
| Dyspnea, n (%) | 7 (17.1) | 19 (52.8) | NA | NA | **0.0015** | | 6 (23.1) | 11 (68.8) | **0.0084** | 1 (33.3) | 2 (100) | 0.4 |
| Odynophagia, n (%) | 21 (51.2) | 6 (16.7) | NA | NA | **0.0019** | | 15 (57.7) | 2 (12.5) | **0.0045** | 1 (33.3) | 0 (0) | 1 |
| ***Constitutional*** | | | | | | | | | | | | |
| Fever, n (%) | 22 (53.7) | 30 (83.3) | NA | NA | **0.0073** | | 14 (53.8) | 14 (87.5) | **0.0420** | 1 (33.3) | 2 (100) | 0.4 |
| Headache, n (%) | 34 (82.9) | 13 (36.1) | NA | NA | **< 0.0001** | | 23 (88.5) | 5 (31.3) | **0.0004** | 2 (66.7) | 2 (100) | 1 |
| Myalgia, n (%) | 26 (63.4) | 17 (47.2) | NA | NA | 0.1742 | | 16 (61.5) | 9 (56.3) | 0.7570 | 3 (100) | 0 (0) | 0.1 |
| Severe fatigue, n (%) | 0 (0) | 21 (58.3) | NA | NA | **< 0.0001** | | 0 (0) | 11 (68.8) | **< 0.0001** | 0 (0) | 1 (50) | 0.4 |
| Altered mental status, n (%) | 0 (0) | 3 (8.3) | NA | NA | 0.0976 | | 0 (0) | 1 (6.3) | 0.3810 | 0 (0) | 0 (0) | NA |

Abbreviations: IQR, interquartile range; NA, not applicable; NS, not statistically significant.
+ Allergy considered self-reported allergic rhinitis (by seasonal, perennial/year-round, or episodic allergens) and food allergy.
Categorical variables compared using Fisher-Freeman-Halton's exact test*; Pairwise Fisher's exact test#; Fisher's exact test^.
Continuous variables compared using Kruskal-Wallis test*; Wilcoxon Rank Sum#.

**Supplementary Table 2. List of reagents and resources used in this study.**

| REAGENT or RESOURCE | SOURCE | IDENTIFIER |
|---|---|---|
| **Anti-Human IgG1-PE** | Southern Biotech | HP6001 |
| **Anti-human IgG2-PE** | Southern Biotech | 31-7-4 |
| **Anti-human IgG3-PE** | Southern Biotech | HP6050 |
| **Anti-human IgG4-PE** | Southern Biotech | HP6025 |
| **Anti-human IgM-PE** | Southern Biotech | SA-DA4 |
| **Anti-human IgA1-PE** | Southern Biotech | HP6025 |
| **Human FcγRIIA** | Duke Human Vaccine Institute | Custom Order |
| **Human FcγRIIB** | Duke Human Vaccine Institute | Custom Order |
| **Human FcγRIIIAV** | Duke Human Vaccine Institute | Custom Order |
| **Human FcγRIIIB** | Duke Human Vaccine Institute | Custom Order |
| **Anti-CD66b Pac Blue** | BioLegend | 305112 |
| **SARS-CoV-2 WT S1** | Sino Biological | 40591-V08H |
| **SARS-CoV-2 WT S2** | Sino Biological | 40590-V08B |
| **SARS-CoV-2 WT Spike** | Sino Biological | 40589-V08H4 |
| **SARS-CoV-2 WT NTD** | Sino Biological | 40591-V49H |
| **SARS-CoV-2 WT Receptor Binding Domain (RBD)** | Sino Biological | 40592-V08H |
| **SARS-CoV-2 Alpha Variant S** | Sino Biological | 40589-V08B6 |
| **SARS-CoV-2 Alpha Variant RBD** | Sino Biological | 40592-V08H82 |
| **SARS-CoV-2 Beta Variant S** | Sino Biological | 40589-V08B7 |
| **SARS-CoV-2 Beta Variant RBD** | Sino Biological | 40592-V08H59 |
| **SARS-CoV-2 Gamma Variant S** | Sino Biological | 40589-V08B10 |
| **SARS-CoV-2 Gamma Variant RBD** | Sino Biological | 40592-V08H86 |
| **SARS-CoV-2 Delta Variant S** | Sino Biological | 40589-V08B16 |
| **SARS-CoV-2 Delta Variant RBD** | Sino Biological | 40592-V08H115 |
| **SARS-CoV-2 Omicron Variant S** | Sino Biological | 40589-V08H26 |
| **SARS-CoV-2 Nucleocapsid** | Sino Biological | 40588-V08B |
| **SARS-CoV-2 Omicron Variant RBD** | Sino Biological | 40592-V08H121 |
| **Human Coronavirus OC43 S** | Sino Biological | 40607-V08B |
| **Human CoV HKU1 S (isolate N5)** | Sino Biological | 40606-V08B |
| **PE-Streptavidin** | Agilent Technologies | PB32-10 |
| **NHS-Sulfo-LC-LC Kit** | ThermoFisher | 21435 |
| **Zebra-Spin Desalting and Chromatography Columns** | ThermoFisher | 89882 |
| **Green Fluorescent Neutravidin Microspheres** | ThermoFisher | F8776 |
| **Red Fluorescent Neutravidin Microspheres** | ThermoFisher | F8775 |

| | | |
|---|---|---|
| **MagPlex Microspheres** | Luminex MFG | MC12001-01 (Cataloged by region) |
| **THP-1 Monocytes** | ATCC | CVCL_0006 |
| **384-well HydroSpeed Plate Washer** | Tecan | 30190112 |
| **iQue Screener Plus** | Intellicyt/Sartorius | 11811 |
| **iQue Forecyt** | Sartorius | 60028 |
| **Vero E6 cells** | ATCC | CRL-1586, RRID: CVCL_0574 |
| **MEM** | Gibco | 11095-080 |
| **FBS** | Gibco | 16000-044 |
| **pCEP4-myc-ACE2** | Addgene | 141185 |
| **Hygromycin** | Invitrogen | 10687010 |
| **Optical bottom 96-well plates** | Thermo Scientific | 165305 |
| **Formaldehyde** | Pierce | 28906 |
| **4′,6-diamidino-2-phenylindole (DAPI)** | Invitrogen | D1306 |
| **R Studio V 1.4.1103** | RStudio, PBC | Open Source |
| **GraphPad Prism** | GraphPad Software, LLC | Ragon Site License |
| **FlowJo V. 10.8** | FlowJo, LLC | www.flowjo.com/solutions/flowjo/downloads |

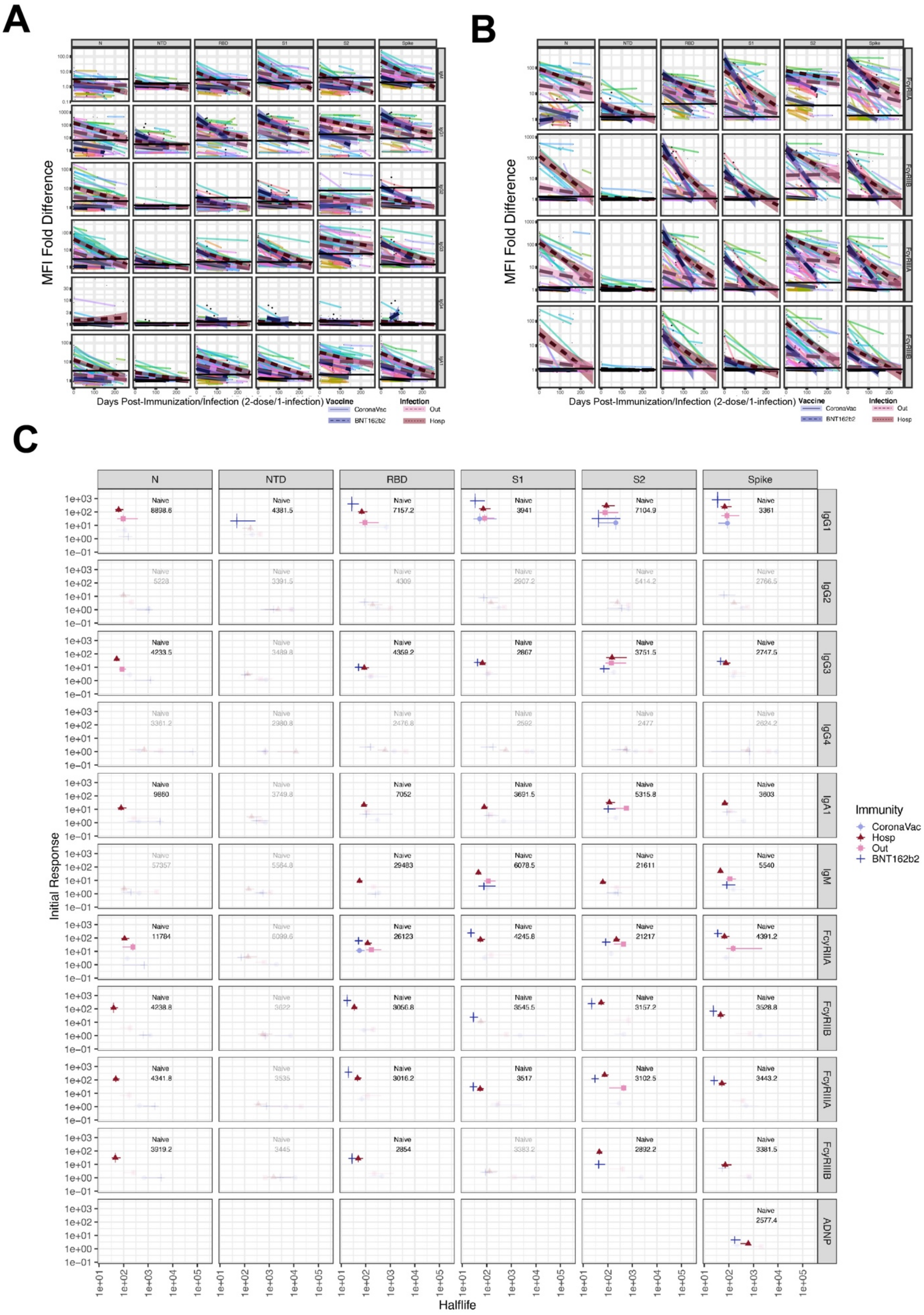
A
B
C
N
NTD
RBD
S1
S2
Spike
MFI Fold Difference
Days Post-Immunization/Infection (2-dose/1-infection)
Vaccine
CoronaVac
BNT162b2
Infection
Out
Hosp
IgG1
IgG2
IgG3
IgG4
IgA1
IgM
FcγRIIA
FcγRIIB
FcγRIIIA
FcγRIIIB
ADNP
Initial Response
Halflife
Immunity
CoronaVac
Hosp
Out
BNT162b2

**Supplemental FigS1. WT Spike subdomain-specific MFI decay over time post-infection or vaccination for Immunoglobulin and FcγR.** (**A, B**) Scatter plots show subject-specific (line color) decline in immunoglobulin response and FcγR binding specific to WT Spike subdomains. Log-linear mixed-effect models with subject-specific random intercepts and slopes estimated trend-lines and 95% confidence intervals using data from subjects following the second BNT162b2 dose (dark-violet), second CoronaVac dose (light-blue), Hospitalized individuals (dark-purple), or Outpatient (light-pink). Horizontal grey lines indicate the innate Spike reactivity in Naïve (no-exposure) samples. Color legend shown at bottom for each panel. (**C**) Regression intercepts and slopes, indicating initial response and decay rate, are plotted with 95% confidence intervals for by each Spike subdomain. Decay and response parameters across variants (shapes) are stratified by infection/vaccination type (color and shape). Shaded out regions indicate a response < the 97.5$^{th}$ percentile of the naïve response. Y-axis units are MFI fold difference respect to the Naïve group.

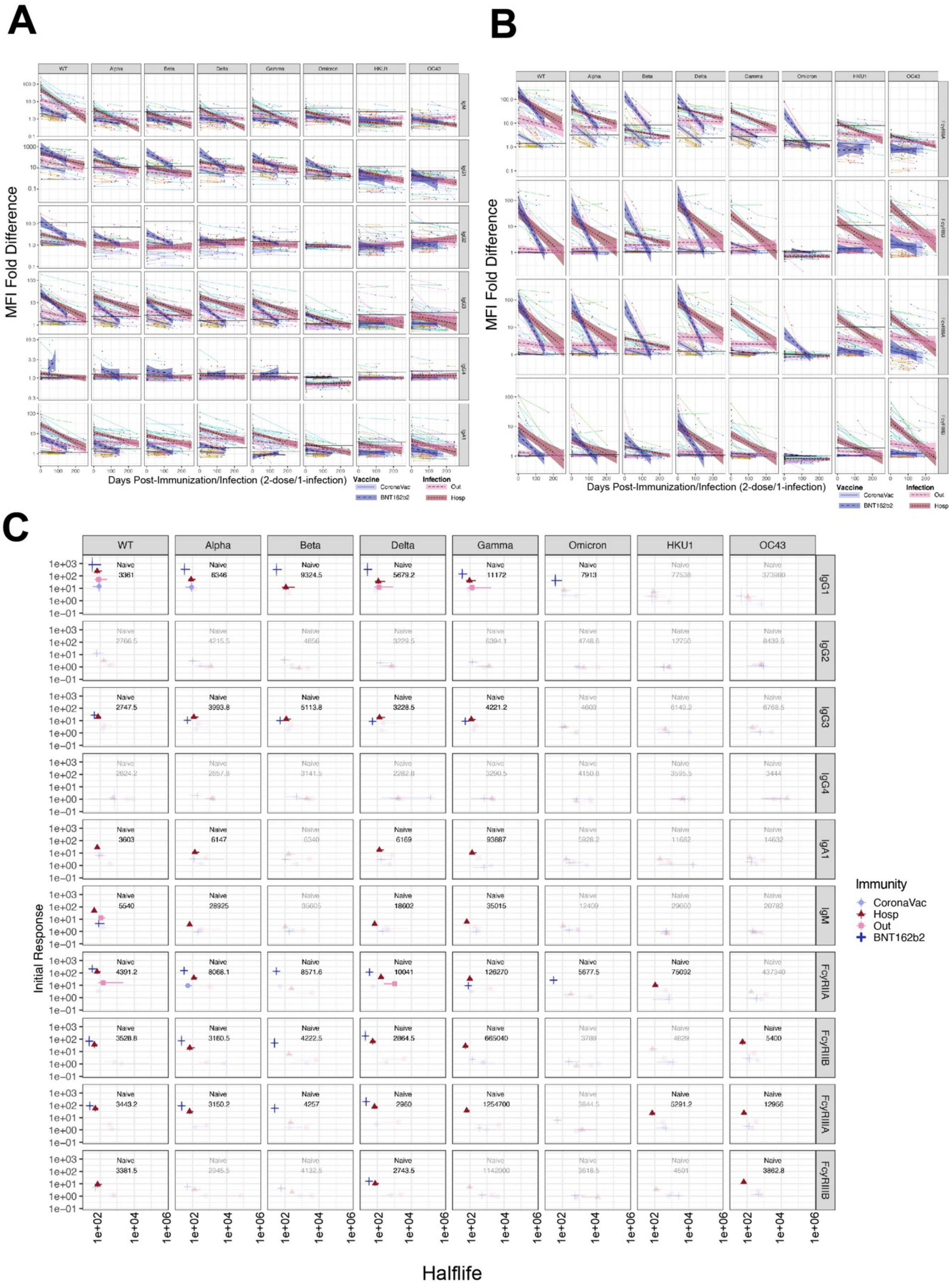
A
B
C
WT
Alpha
Beta
Delta
Gamma
Omicron
HKU1
OC43
MFI Fold Difference
Days Post-Immunization/Infection (2-dose/1-infection)
Vaccine
CoronaVac
BNT162b2
Infection
Out
Hosp
IgG1
IgG2
IgG3
IgG4
IgA1
IgM
FcγRIIA
FcγRIIB
FcγRIIIA
FcγRIIIB
Initial Response
Halflife
Immunity
CoronaVac
Hosp
Out
BNT162b2

**Supplemental FigS2. Spike-specific MFI decay across VOC over time post-infection or vaccination for Immunoglobulin and FcR.** (**A, B**) Scatter plots show subject-specific (line color) decline in immunoglobulin response and FcR binding specific to various VOC Spikes. Log-linear mixed-effect models with subject-specific random intercepts and slopes estimated trend-lines and 95% confidence intervals using data from subjects following the second BNT162b2 (dark-violet), dose, second CoronaVac dose (light-blue), Hospitalized individuals (dark-purple), or Outpatient (light-pink). Horizontal grey lines indicate the innate Spike reactivity in Naïve (no-exposure) samples. Color legend shown at bottom for each panel. (**C**) Regression intercepts and slopes, indicating initial response and decay rate, are plotted with 95% confidence intervals for by each Spike subdomain. Decay and response parameters across variants (shapes) are stratified by infection/vaccination type (color and shape). Shaded out regions indicate a response < the 97.5$^{th}$ percentile of the naïve response. Y-axis units are MFI fold difference respect to the Naïve group.

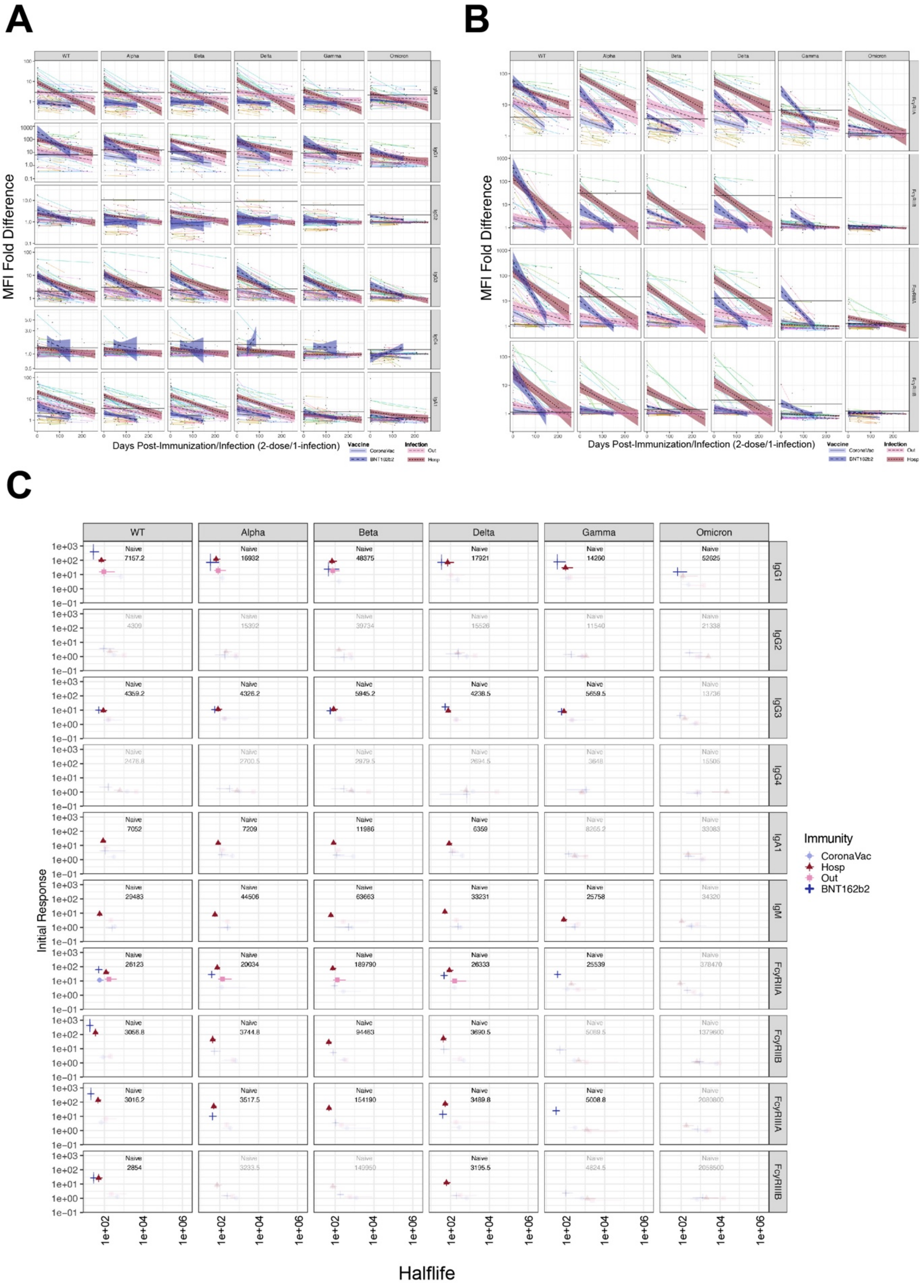
A
B
C
MFI Fold Difference
Days Post-Immunization/Infection (2-dose/1-infection)
Vaccine
CoronaVac
BNT162b2
Infection
Out
Hosp
WT
Alpha
Beta
Delta
Gamma
Omicron
IgG1
IgG2
IgG3
IgG4
IgA1
IgM
FcγRIIA
FcγRIIB
FcγRIIIA
FcγRIIIB
Initial Response
Halflife
Immunity
CoronaVac
Hosp
Out
BNT162b2

**Supplemental FigS3. RBD-specific MFI decay across VOC over time post-infection or vaccination for Immunoglobulin and FcR.** (**A,B**) Scatter plots show subject-specific (line color) decline in immunoglobulin response and FcR binding specific to various VOC RBDs. Log-linear mixed-effect models with subject-specific random intercepts and slopes estimated trend-lines and 95% confidence intervals using data from subjects following the second BNT162b2 (dark-violet), dose, second CoronaVac dose (light-blue), Hospitalized individuals (dark-purple), or Outpatient (light-pink). Horizontal grey lines indicate the innate spike reactivity in Naïve (no-exposure) samples. Color legend shown at bottom for each panel. (**C**) Regression intercepts and slopes, indicating initial response and decay rate, are plotted with 95% confidence intervals for by each Spike subdomain. Decay and response parameters across variants (shapes) are stratified by infection/vaccination type (color and shape). Shaded out regions indicate a response < the 97.5$^{th}$ percentile of the naïve response. Y-axis units are MFI fold difference respect to the Naïve group.

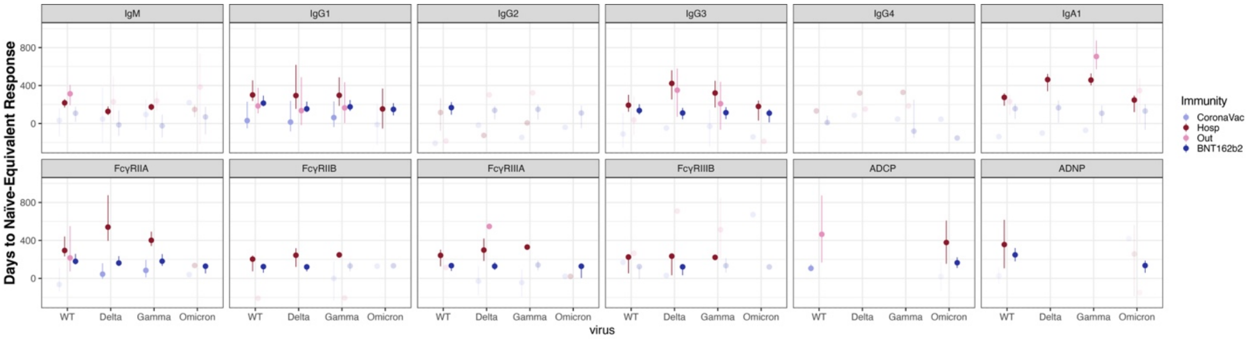


**Supplemental FigS4. Time to decay calculated from the log-linear regression models.** Point-ranges display the predicted decay periods over which each inoculation- or infection-induced initial response to corresponding naïve (no-exposure) response; the range corresponds to the time predicted for each model to reach the 97.5th, 50th, and 2.5th percentile of the naïve response distribution s.t. Time = (Naïve$_{97.5th,\ 50th,\ 2.5th}$ – Decay)/Initial. Color legend for group is shown on the right.

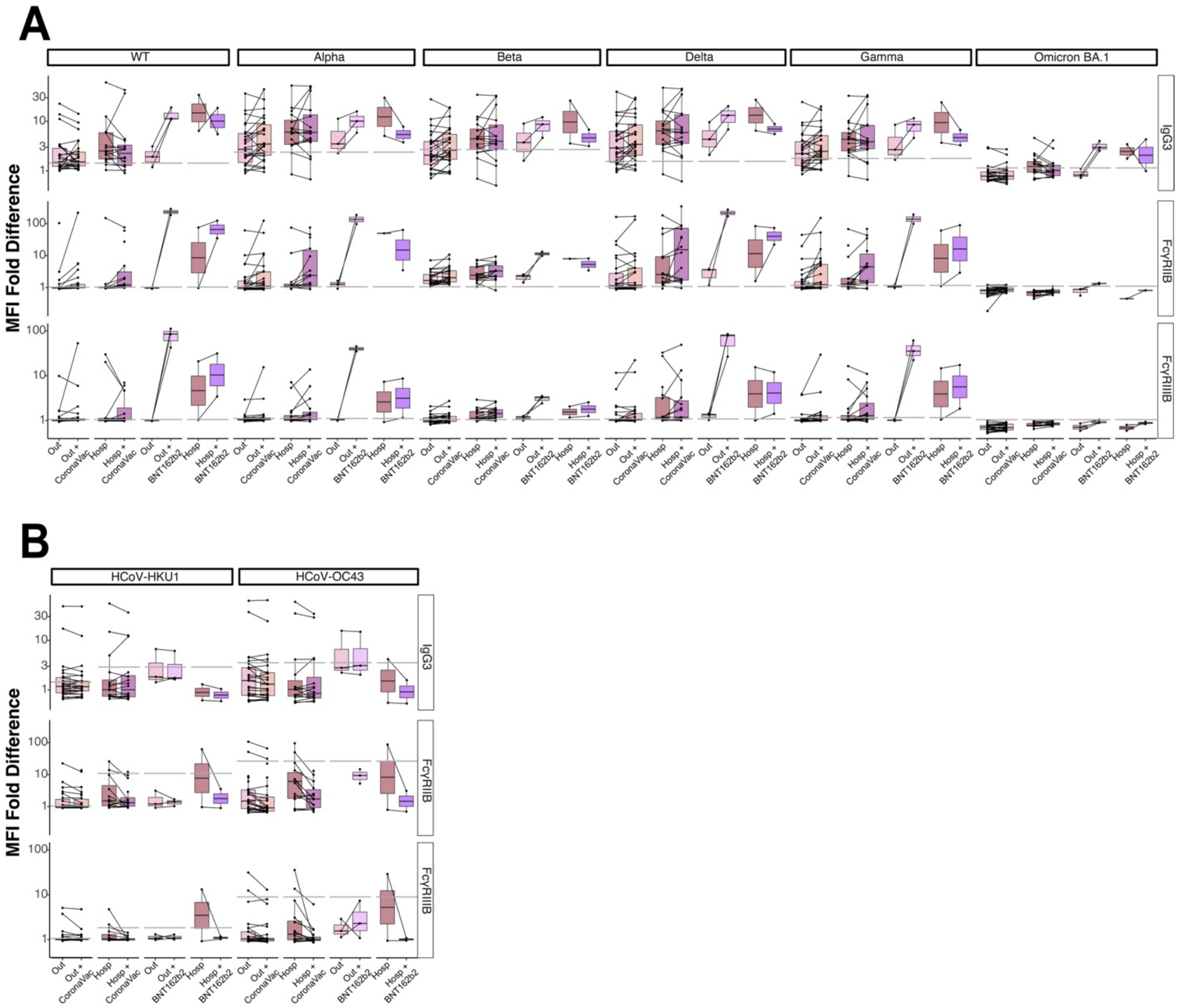


**Supplemental FigS5. Comparison of the peak response post-infection to the peak response following post-infection across VOC for Immunoglobulin and FcR.** Paired t-tests are visualized for various VOC Spikes binding (**A, B**) for naturally infected individuals (hospitalized or outpatients) immunized with either the CoronaVac or BNT162b2 vaccines post-infection. The infection-only boxplots (Out or Hosp) represent the fully waned response (response at the farthest timepoint following infection) while the paired post-infection vaccination boxplots represent peak immunity following vaccination. The grey line indicates the 97th percentile of naïve response. Q-values indicate the FDR corrected significance for each corresponding paired t-test. Y-axis units for A and B are MFI fold difference respect to the Naïve group.

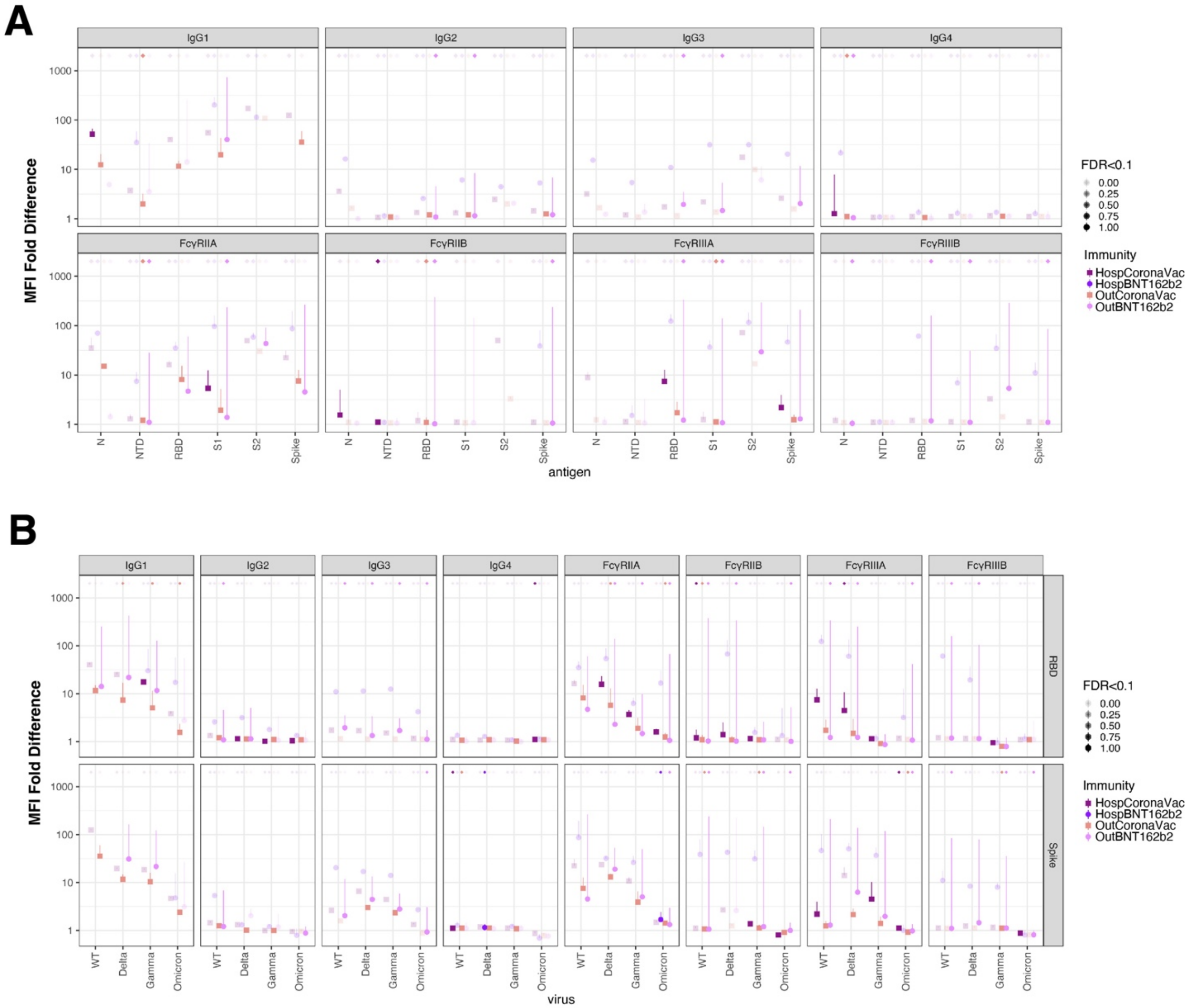


**Supplemental FigS6. Hybrid boosting expands binding breadth for both outpatient and hospitalized recoverees**. Point-ranges indicating the lowest response observed post-infection (circle/square) and the increase in response observed post-vaccination (line height) where response is log10(MFI/MFI_naive). Plots are stratified by assay, antigen, and VOC and colored by infection and vaccination type. Point-ranges are transparent if a multiple test corrected paired t-test on response is not significant (FDR>0.1). An opaque diamond is added above each point-range if the waned response is less than naïve, the boosted response is greater than naïve, and the multiple test corrected paired t-test is significant. Color legend for group is shown on the right. Y-axis units are MFI fold difference.

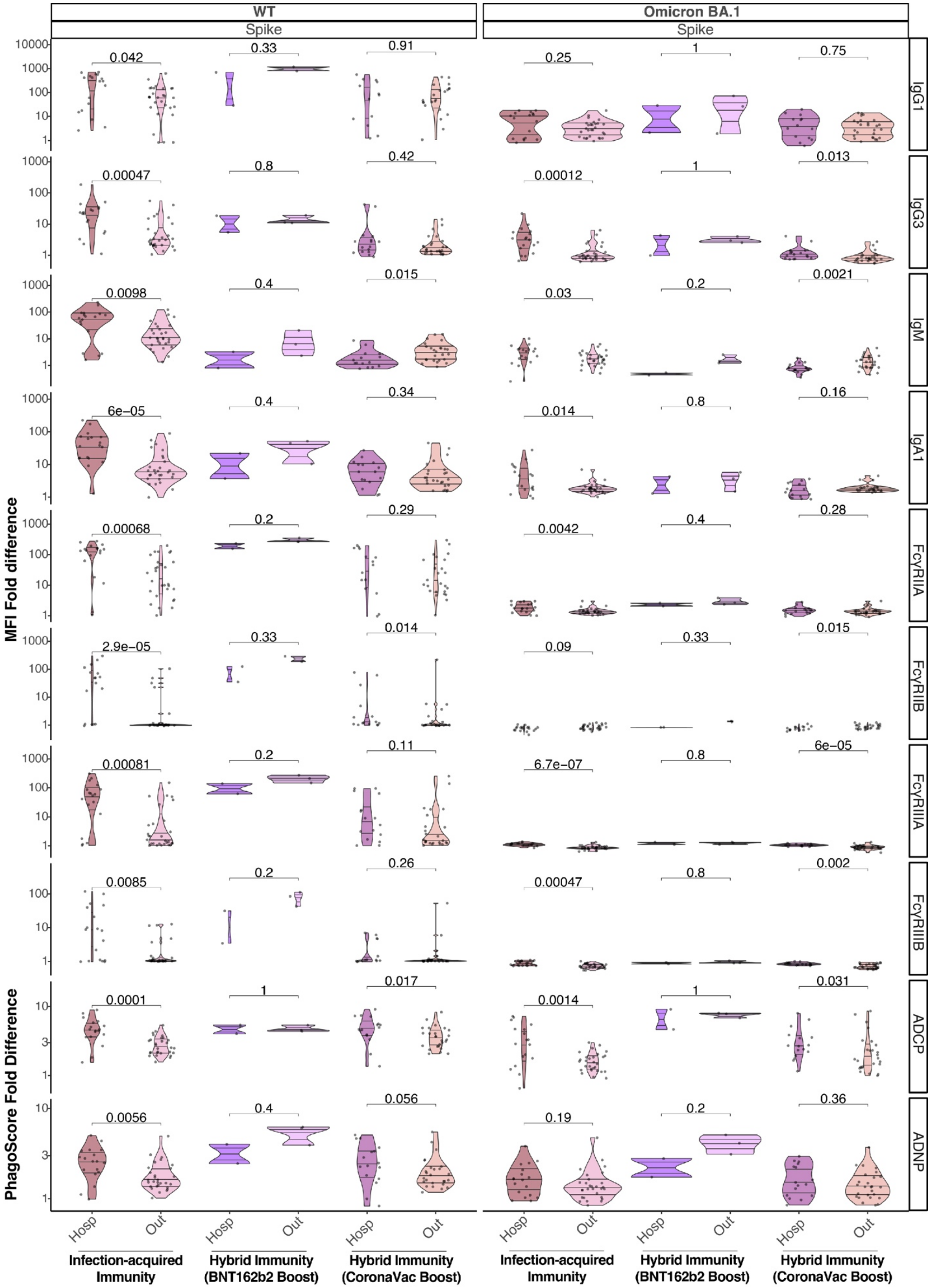

WT
Omicron BA.1
Spike
Spike
IgG1
IgG3
IgM
IgA1
FcγRIIA
FcγRIIB
FcγRIIIA
FcγRIIIB
ADCP
ADNP
MFI Fold difference
PhagoScore Fold Difference
0.042
0.33
0.91
0.25
1
0.75
0.00047
0.8
0.42
0.00012
1
0.013
0.0098
0.4
0.015
0.03
0.2
0.0021
6e−05
0.4
0.34
0.014
0.8
0.16
0.00068
0.2
0.29
0.0042
0.4
0.28
2.9e−05
0.33
0.014
0.09
0.33
0.015
0.00081
0.2
0.11
6.7e−07
0.8
6e−05
0.0085
0.2
0.26
0.00047
0.8
0.002
0.0001
1
0.017
0.0014
1
0.031
0.0056
0.4
0.056
0.19
0.2
0.36
Hosp
Out
Infection-acquired Immunity
Hybrid Immunity (BNT162b2 Boost)
Hybrid Immunity (CoronaVac Boost)

**Supplemental FigS7. Comparison of the peak response post-infection to the peak response following post-infection.** Violin plots comparing the immune response (MFI/MFI_naive) to Omicron and WT Spike binding and function given either CoronaVac or Pfizer vaccination post-infection. (a) Violin plots compare the peak response post-infection to the peak response following post-infection vaccination. Q-values indicate the FDR corrected significance for each corresponding t-test.

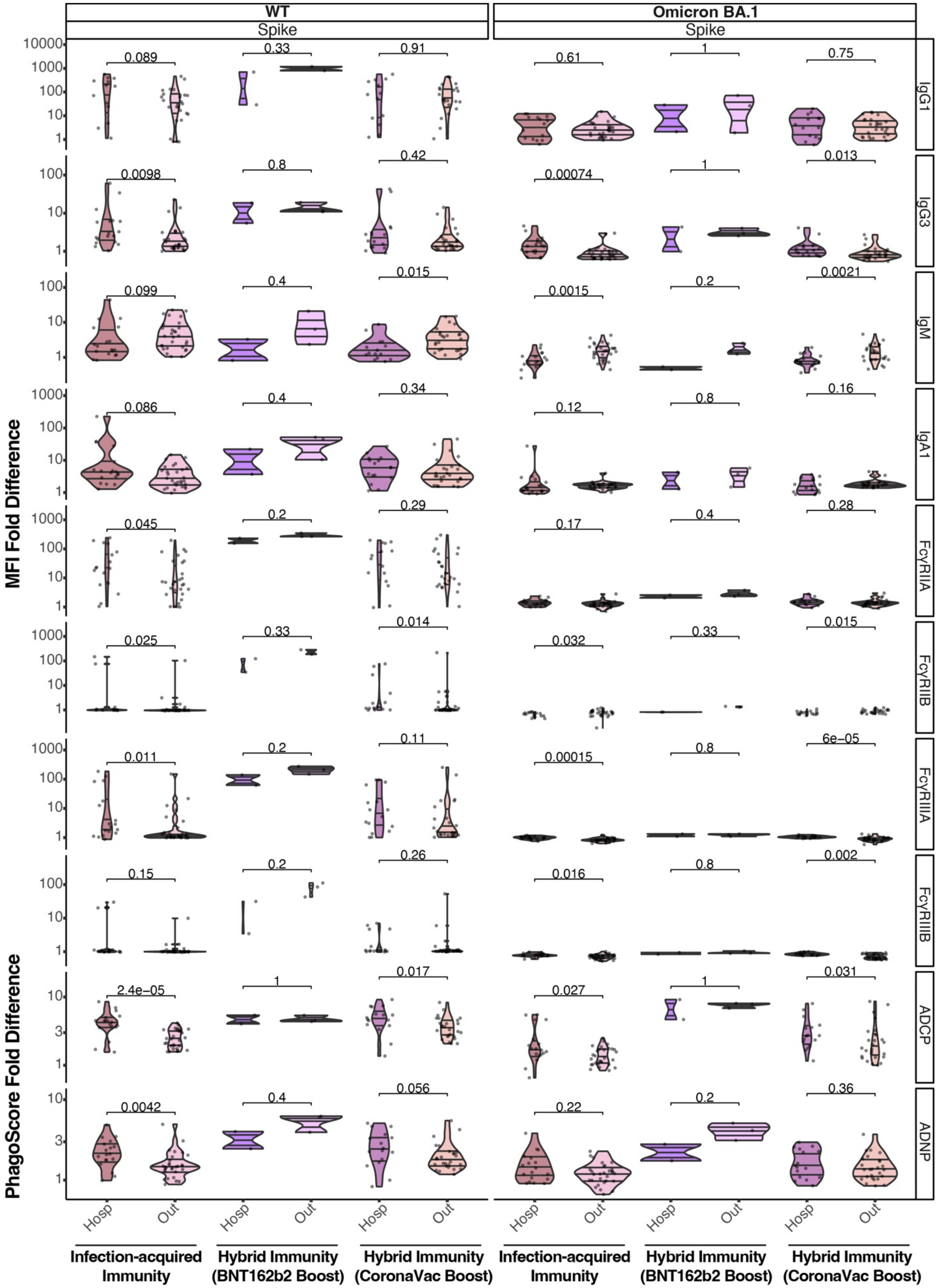
WT
Spike
Omicron BA.1
Spike
MFI Fold Difference
PhagoScore Fold Difference
IgG1
IgG3
IgM
IgA1
FcγRIIA
FcγRIIB
FcγRIIIA
FcγRIIIB
ADCP
ADNP
0.089
0.33
0.91
0.61
1
0.75
0.0098
0.8
0.42
0.00074
1
0.013
0.099
0.4
0.015
0.0015
0.2
0.0021
0.086
0.4
0.34
0.12
0.8
0.16
0.045
0.2
0.29
0.17
0.4
0.28
0.025
0.33
0.014
0.032
0.33
0.015
0.011
0.2
0.11
0.00015
0.8
6e−05
0.15
0.2
0.26
0.016
0.8
0.002
2.4e−05
1
0.017
0.027
1
0.031
0.0042
0.4
0.056
0.22
0.2
0.36
Hosp
Out
Infection-acquired Immunity
Hybrid Immunity (BNT162b2 Boost)
Hybrid Immunity (CoronaVac Boost)

**Supplemental FigS8. Comparison of waning with peak response following post-infection boosting.** Violin plots comparing the immune response (MFI/MFI_naive) to Omicron and WT Spike binding and function given either CoronaVac or Pfizer vaccination post-infection. Violin plots compare waning with peak response following post-infection boosting. The infection-only boxplots (Out or Hosp) represent the fully waned response (response at the farthest timepoint following infection) while the paired post-infection vaccination boxplots represent peak immunity following vaccination. Q-values indicate the FDR corrected significance for each corresponding t-test.
